# The tryptophan catabolite or kynurenine pathway in Long COVID disease: A systematic review and meta-analysis

**DOI:** 10.1101/2024.04.30.24306635

**Authors:** Abbas F. Almulla, Yanin Thipakorn, Bo Zhou, Aristo Vojdani, Rossitsa Paunova, Michael Maes

## Abstract

**Background:** Recent studies confirm the involvement of activated immune-inflammatory responses and increased oxidative and nitrosative stress in Long COVID (LC) disease. However, the influence of these pathways on the metabolism of tryptophan (TRP) through the TRP catabolite (TRYCAT) pathway and their mediating effects on LC pathophysiology, has not been fully explored.

**Objective:** This meta-analysis investigates peripheral TRP and TRYCAT levels and the TRYCAT pathway in patients with LC disease.

**Method:** This review utilized systematic searches of PubMed, Google Scholar, and SciFinder, including 14 full-text articles and 1,167 participants, consisting of 480 patients with LC and 687 normal controls.

**Results:** The results indicated a significant increase in the kynurenine (KYN)/TRP ratio, with a large effect size (standardized mean difference, SMD = 0.755; confidence intervals, CI: 0.119;1.392), in LC patients compared to normal controls. Additionally, LC patients exhibited a significant decrease in TRP levels (SMD = -0.520, CI: -0.793; -0.246) and an increase in KYN levels after imputing missing studies (SMD = 1.176, CI: 0.474; 1.877), suggesting activation of the Indoleamine 2,3-dioxygenase (IDO) enzyme and upregulation of the TRYCAT pathway. No significant elevation in TRYCAT-related neurotoxicity, kynurenic acid (KA)/KYN and 3-hydroxykynurenine (3-HK)/KYN ratios were observed in LC patients compared to normal controls.

**Conclusion:** The current findings indicate that an activated TRYCAT pathway, characterized by decreased TRP levels and maybe elevated KYN levels, plays a significant role in the pathophysiology of LC.

## Introduction

The severe acute respiratory syndrome coronavirus 2 (SARS-CoV-2) pandemic has precipitated not only the acute manifestations of Coronavirus disease 2019 (COVID-19) but also the emergence of Long COVID (LC) (NICE 2020). This syndrome is characterized by enduring symptoms that persist or emerge following the resolution of the initial infection (Phillips and Williams 2021) which impact the quality of life of recovered individuals (Maes, Al-Rubaye et al. 2022). Commonly reported symptoms include, but are not limited to, fatigue and dyspnoea, distress symptoms, and cognitive impairments such as issues with concentration and memory. Mood disorders including anxiety and depression are also prevalent among affected individuals (Nalbandian, Sehgal et al. 2021, Badenoch, Rengasamy et al. 2022, Ceban, Ling et al. 2022).

In previous work, we identified one validated latent construct termed the ’physio-affective phenome’ that underpinned the affective and physiosomatic (previously labeled as psychosomatic symptoms) both in the acute infectious phase and in LC. This construct, derived from physiosomatic symptoms, including chronic fatigue syndrome (CFS) and fibromyalgia, depressive and anxiety symptoms, has demonstrated that these manifestations share a common underlying core, which is pivotal in understanding the full impact of the disease (Al-Hadrawi, Al-Rubaye et al. 2022, Al-Hakeim, Al-Rubaye et al. 2022, Al-Jassas, Al-Hakeim et al. 2022, Al-Hakeim, Al-Rubaye et al. 2023, Al-Hakeim, Khairi Abed et al. 2023). Our observations link various biological anomalies to the physio-affective phenome of LC patients (Almulla, Al-Hakeim et al. 2023). Notably, significant associations between the diagnosis LC and the physio-affective phenome due to LC and activated immune-inflammatory pathways, as well as oxidative and nitrosative stress were established (Al-Hakeim, Al-Rubaye et al. 2022, Al-Hakeim, Al-Rubaye et al. 2023, Al-Hakeim, Al-Rubaye et al. 2023). It is critical to recognize that these aberrant pathways may at least partly result from the acute phase of the illness, where elevated peak body temperature (PBT) and reduced oxygen saturation (SpO2) could account for significant variations in these pathways (Al-Hakeim, Al-Rubaye et al. 2022, Al-Hakeim, Al-Rubaye et al. 2023, Al-Hakeim, Al-Rubaye et al. 2023). As explained previously, both increased PBT and lowered SpO2 are markers of the inflammatory response during acute COVID-19 (Al-Hakeim, Al-Rubaye et al. 2022, Al-Hakeim, Al-Rubaye et al. 2023, Al-Hakeim, Al-Rubaye et al. 2023).

In a recent extensive meta-analysis, we discovered that LC is characterized by an activated immune-inflammatory response (IRS), including activation of M1 macrophages and T cells, as well as immune-related neurotoxicity (Abbas, Yanin et al. 2024). Thus, significant elevations in several inflammatory markers were observed, such as C-reactive protein (CRP) and various cytokines/chemokines and growth factors (Abbas, Yanin et al. 2024). These immune mediators and reactive oxygen and nitrogen species (RONS) notably induce the activity of indoleamine 2,3-dioxygenase (IDO), the rate-limiting enzyme in the TRP catabolic pathway (Maes, Leonard et al. 2011, Maes 2015, Almulla and Maes 2022). This activation results in reduced blood levels of TRP and its neuroprotective derivatives such as serotonin and melatonin and increases in neuroactive metabolites (Almulla and Maes 2022, Almulla and Maes 2023).

In patients with acute and severe/critical COVID-19, we observed a significantly elevated KYN/TRP ratio, indicative of an activated IDO enzyme and, consequently, upregulation of the TRYCAT pathway. This was accompanied by decreased levels of TRP and increased levels of KYN (Almulla, Supasitthumrong et al. 2022). Similar patterns were recently observed in LC patients, where these markers significantly correlated with the physio-affective phenome of LC (Al-Hakeim, Abed et al. 2023, Al-Hakeim, Khairi Abed et al. 2023). Earlier studies have also reported a significant activation of the TRYCAT pathway with reduced TRP levels in LC, supporting the role of this pathway in the persistent degradation of TRP and increased TRP metabolites in LC disease (Jud, Gressenberger et al. 2021, Piater, Gietl et al. 2023, Krčmová, Javorská et al. 2024).

No prior meta-analyses have investigated TRP levels and the TRYCAT pathway in LC. Thus, our current study aims to evaluate TRP levels alongside the activities of enzymes such as IDO, kynurenine aminotransferase (KAT), and kynurenine 3-monooxygenase (KMO), using ratios like KYN/TRP, KA/KYN, and 3-HK/KYN as proxies. We will also assess levels of TRYCATs including KYN, KA, and 3-HK in LC patients compared to controls, hypothesizing that LC is associated with persistently activated TRYCAT pathway and lowered TRP levels.

## Materials and method

Our study focused on evaluating peripheral TRP, and its catabolites including KYN, KA, and 3-HK in patients diagnosed with LC disease versus normal controls. Additionally, we assessed composite indices for IDO, KAT, and KMO enzymes activities along with TRYCAT-related neurotoxicity index by analyzing combined neurotoxic metabolites, such as KYN, 3-hydroxy anthranilic acid (3-HAA), xanthurenic acid (XA), quinolinic acid (QA), and picolinic acid (PA). To ensure a thorough and methodologically sound approach, we employed several key frameworks, including the Preferred Reporting Items for Systematic Reviews and Meta-Analyses (PRISMA) 2020 (Page, McKenzie et al. 2021), the Cochrane Handbook for Systematic Reviews of Interventions (Higgins JPT 2019), and the guidelines for conducting Meta-Analyses of Observational Studies in Epidemiology.

### Search Strategy

Our extensive literature search aimed at collecting data on TRP and its catabolites in LC spanned from January to March 15th, 2024, and included a thorough examination of databases such as PubMed/MEDLINE, Google Scholar, and SciFinder. By employing a targeted selection of keywords and MeSH terms, as outlined in Table 1 of the supplementary electronic file (ESF), we also meticulously reviewed reference lists from pertinent studies to ensure no relevant study was overlooked, thereby achieving comprehensive literature coverage.

**Table 1.** The outcomes and number of patients with Long COVID and normal controls along with the side of standardized mean difference (SMD) and the 95% confidence intervals with respect to zero SMD.

| Outcome profiles | n studies | Side of 95% confidence intervals |  |  |  | Patient Cases | Control Cases | Total number of participants |
| --- | --- | --- | --- | --- | --- | --- | --- | --- |
|  |  | < 0 | Overlap 0 and SMD < 0 | Overlap 0 and SMD > 0 | > 0 |  |  |  |
| KYN/TRP | 14 | 0 | 1 | 6 | 7 | 480 | 687 | 1167 |
| TRP | 10 | 3 | 6 | 1 | 0 | 307 | 292 | 599 |
| KYN | 11 | 0 | 2 | 4 | 5 | 411 | 603 | 1014 |
| Neurotoxicity | 11 | 0 | 2 | 5 | 4 | 411 | 603 | 1014 |
| KA/KYN | 3 | 0 | 2 | 1 | 0 | 134 | 104 | 238 |
| 3HK/KYN | 3 | 2 | 0 | 1 | 0 | 134 | 104 | 238 |
| KA | 3 | 1 | 1 | 0 | 1 | 134 | 104 | 238 |
| 3-HK | 3 | 2 | 0 | 1 | 0 | 134 | 104 | 238 |

KYN kynurenine, TRP tryptophan, KA kynurenic acid, 3-HK: 3-Hydroxykynurenine

### Eligibility criteria

In this meta-analysis, we primarily assessed articles in English from peer-reviewed journals, expanding our scope to include grey literature and studies published in different languages including Thai, French, Spanish, Turkish, German, Italian, and Arabic. Our focus was on observational studies—either cohort or case-control—that investigated the levels of TRP and its metabolites, such as KYN and KA, among others. We ensured that the included patients with Long or Post COVID disease met the World Health Organization (WHO) diagnostic criteria (World Health Organization 2021). Additionally, we incorporated studies that provided baseline assessments of TRP and its catabolites and their follow-up evaluations. Exclusion criteria applied to studies utilizing unconventional media (e.g., hair, saliva, platelet-rich plasma), those without a control group, or lacking mean and standard deviation (SD) or standard error (SE) data for biomarkers. Efforts were made to obtain missing data from authors. Where no response was received, we calculated mean (SD) from the available information using methods from Wan et al. (2014), the online Mean Variance Estimation tool, and resorted to Web Plot Digitizer for extracting mean (SD) from graphical representations (https://automeris.io/WebPlotDigitizer/).

### Primary and secondary outcomes

The primary outcomes of this meta-analysis were the proxy for IDO enzyme activity, as indicated by the KYN/TRP ratio, along with the levels of TRP, KYN, and neurotoxicity indices, which include sum of neurotoxic TRYCATs such as KYN, 3-HAA3-HK, QA, XA, and PA in LC patients compared to controls, as outlined in **Table 1**. Secondary outcomes were the KA/KYN and 3-HK/KYN ratios as markers of KAT and KMO enzyme activities, respectively, in addition to measuring the levels of other TRYCATs, specifically KA and 3HK.

### Screening and data extraction

In the initial phase, AA and YT undertook the screening of studies by reviewing titles and abstracts against the inclusion criteria to determine the eligibility for our meta-analysis. Subsequent steps involved the acquisition and evaluation of full-text articles, discarding those that did not align with our criteria. This meticulous selection process facilitated the extraction of essential data into a structured predefined Excel template, capturing details such as mean, SD, and other relevant information from the studies that met our criteria. The Excel template was designed to record comprehensive details, including authors’ names, publication dates, means and SD of TRP and TRYCATs, sample sizes of the patient and control groups, research designs, types of samples (including serum, plasma, CSF, and brain tissues), timing post-acute COVID infection, numbers of patients hospitalized or admitted to the ICU during the acute phase, and participants’ demographic information such as age, gender, and study location. The same two authors conducted a thorough review of the final dataset compiled in the predefined Excel file, resolving any discrepancies through discussions with MM.

The methodological quality of the included studies was assessed using an adapted version of the Immunological Confounder Scale (ICS), as outlined by Andrés-Rodríguez et al., 2020 (Andres-Rodriguez, Borras et al. 2020), and modified by the senior author (MM) for compatibility with TRYCATs research in LC. The adaptation involved a detailed assessment framework comprising quality and “redpoints” scales, extensively described in the ESF (Table 2) and previously utilized by Almulla et al. for evaluating studies on TRYCATs levels in acute COVID-19 (Almulla, Supasitthumrong et al. 2022), autism (Almulla, Thipakorn et al. 2023), Alzheimer’s disease (Almulla, Supasitthumrong et al. 2022), and affective disorders (Almulla, Thipakorn et al. 2022b). The quality scale, ranging from 0 (better quality) to 10, emphasized the importance of sample size, the control of confounders, and sampling duration. Conversely, the “redpoints” scale aimed at predicting potential biases in TRYCATs outcomes and study designs by assessing control over key confounders, with scores ranging from zero (optimal control) to twenty-six (inadequate consideration of confounders).

**Table 2.** Results of meta-analysis performed on several outcome (TRYCATs) variables with combined different media.

| Outcome feature sets | n | Groups | SMD | 95% CI | z | p | Q | df | p | I<sup>2</sup> (%) | $\tau^2$ | T |
| --- | --- | --- | --- | --- | --- | --- | --- | --- | --- | --- | --- | --- |
| KYN/TRP | 14 | Overall | 0.755 | 0.119;1.392 | 2.328 | 0.020 | 263.020 | 13 | 0.000 | 95.057 | 1.370 | 1.170 |
| TRP | 10 | Overall | -0.520 | -0.793; -0.246 | -3.722 | 0.000 | 19.895 | 9 | 0.019 | 54.762 | 0.097 | 0.311 |
| KYN | 11 | Overall | 0.701 | -0.124;1.527 | 1.665 | 0.096 | 280.137 | 10 | 0.000 | 96.430 | 1.844 | 1.358 |
| Neurotoxicity | 11 | Overall | 0.721 | -0.110; 1.552 | 1.700 | 0.089 | 278.186 | 10 | 0.000 | 96.405 | 1.867 | 1.366 |
| KA/KYN | 3 | Overall | 0.028 | -0.240; 0.296 | 0.202 | 0.840 | 2.021 | 2 | 0.364 | 1.060 | 0.001 | 0.027 |
| 3HK/KYN | 3 | Overall | -0.427 | -0.934; 0.080 | -1.651 | 0.099 | 5.241 | 2 | 0.073 | 61.837 | 0.121 | 0.347 |
| KA | 3 | Overall | -0.141 | -1.208;0.925 | -0.260 | 0.795 | 24.878 | 2 | 0.000 | 91.961 | 0.805 | 0.897 |
| 3-HK | 3 | Overall | -0.921 | -2.099;0.256 | -1.534 | 0.125 | 26.583 | 2 | 0.000 | 92.476 | 0.968 | 0.984 |

KYN kynurenine, TRP tryptophan, KA kynurenic acid, 3-HK: 3-Hydroxykynurenine

### Data analysis

This meta-analysis adhered to PRISMA guidelines and employed Comprehensive Meta-Analysis (CMA) V4 software. Inclusion criteria required a minimum of three studies per TRYCAT of interest. Mean outcome values were compared between LC patients and controls. KYN/TRP, KA/KYN, and 3HK/KYN ratios served as proxies for IDO, KAT, and KMO enzyme activity, respectively (Almulla, Vasupanrajit et al. 2022, Almulla, Thipakorn et al. 2022a). While assuming dependence, we selected positive effect size directionality indicated increased KYN and decreased TRP (IDO), positive KA and negative KYN (KAT), and positive 3HK and negative KYN (KMO). The random-effects model with constrained maximum likelihood was chosen due to anticipated participant heterogeneity. Standardized mean difference (SMD) with 95% confidence intervals (CI) was used as the effect size measure, with p-values < 0.05 considered statistically significant. Established criteria (Cohen 2013) defined SMD interpretation (large: ≥0.80, moderate: 0.50-0.79, small: ≤0.49). Heterogeneity was assessed using τ², Q statistic, and I² (Higgins, Thompson et al. 2003).

Meta-regression and subgroup analyses explored potential heterogeneity sources, including media (serum, plasma), sample sizes, and timing after acute COVID infection. Leave-one-out sensitivity analyses evaluated effect size and heterogeneity robustness. Publication bias was assessed using fail-safe N, continuity-corrected Kendall tau, and Egger’s regression intercept tests. Duval and Tweedie’s trim-and-fill method addressed potential publication bias by imputing missing studies and adjusting effect sizes (Duval and Tweedie 2000a, Duval and Tweedie 2000b). Funnel plots visualized study precision versus SMD, incorporating observed and imputed data.”

## Results

### 3.1. Search results

Initially, 103 articles were scrutinized using the Mesh terms and keywords outlined in the ESF, Table 1. The PRISMA flow chart, illustrating the inclusion and exclusion of articles, is shown in Figure 1. After refining the search results, eleven studies deemed redundant or irrelevant were removed. Subsequently, 14 full-text articles that satisfied the predefined inclusion and exclusion criteria were selected for this systematic review. Consequently, the meta-analysis ultimately included 14 studies and examined TRP and TRYCATs among 1,167 participants, consisting of 480 patients with LC and 687 normal controls. (Holmes, Wist et al. 2021, Jud, Gressenberger et al. 2021, Bizjak, Stangl et al. 2022, Guntur, Nemkov et al. 2022, Kucukkarapinar, Yay-Pence et al. 2022, Al-Hakeim, Abed et al. 2023, Al-Hakeim, Khairi Abed et al. 2023, Gietl, Burkert et al. 2023, Guo, Appelman et al. 2023, Kovarik, Bileck et al. 2023, López-Hernández, Monárrez-Espino et al. 2023, Piater, Gietl et al. 2023, Krčmová, Javorská et al. 2024, Saito, Shahbaz et al. 2024).

**Figure 1:**
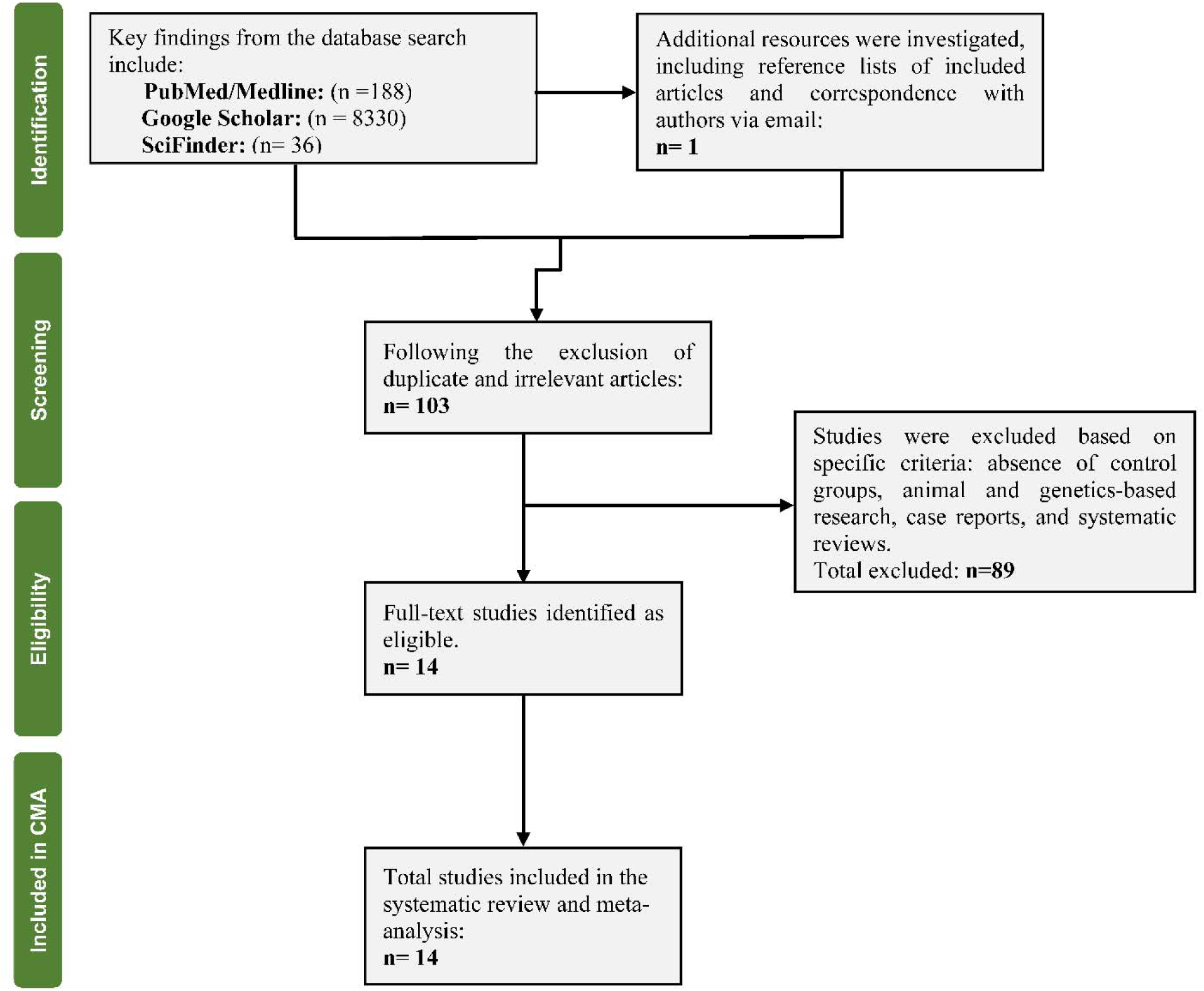
The PRISMA flow chart.

### Primary Outcome Variables

#### KYN/TRP ratio

Fourteen studies contributed data on the KYN/TRP ratio. As summarized in **Table 1**, while no study reported CIs completely below zero, seven studies showed CIs entirely above zero. Another seven studies had CIs that intersected with zero, with one study reporting SMD below zero and 6 studies reporting SMDs above zero. As depicted in **Table 2** and **Figure 2**, a significant increase in the KYN/TRP ratio was observed in LC patients compared to the control group. A publication bias was identified, with 6 studies missing on the right side of the funnel plot (**Table 3**). After adjusting for these missing studies, the effect size increased to 1.141, underscoring a significant rise.

**Figure 2:**
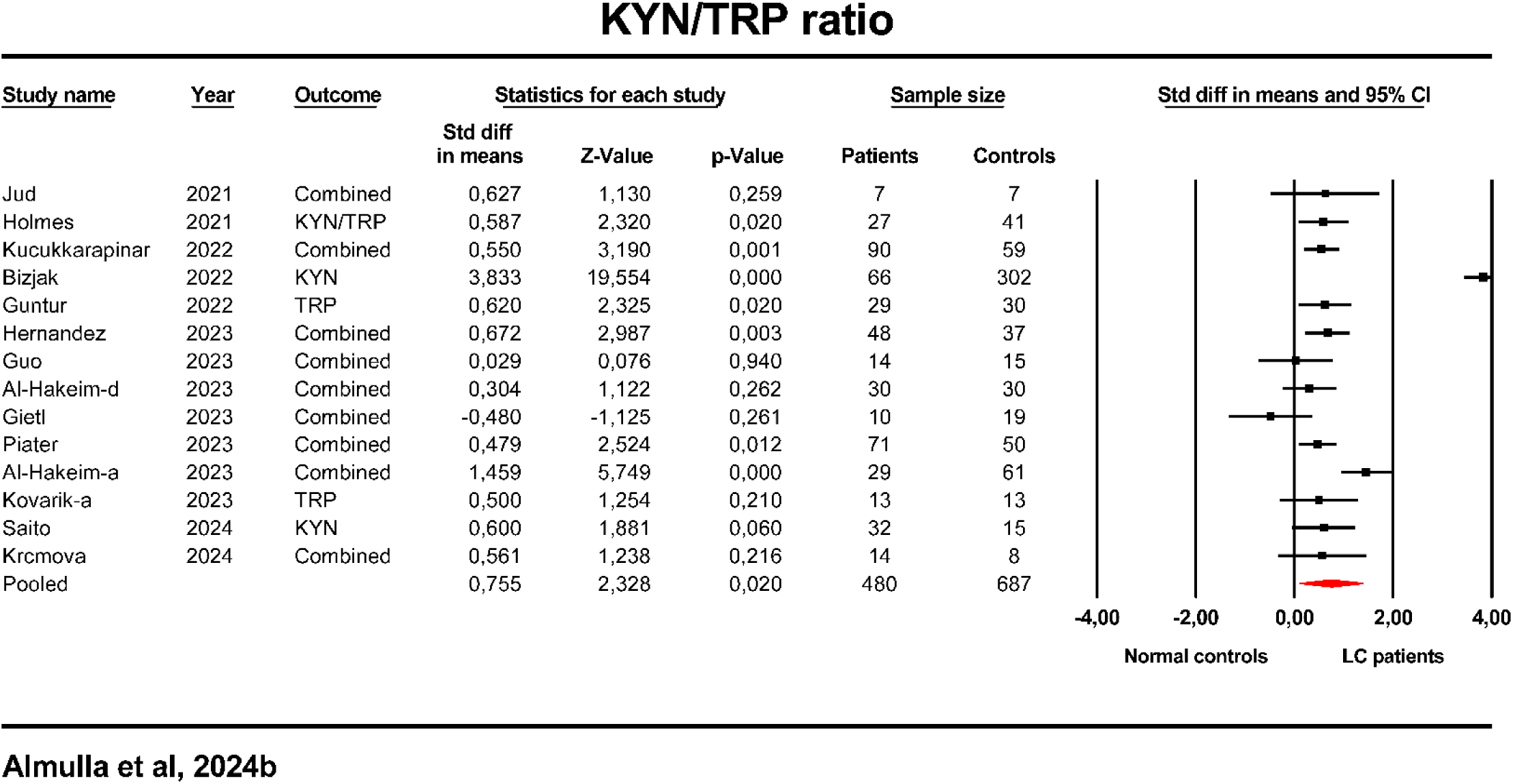
Forest plot with the results of a meta-analysis performed on the kynurenine/tryptophan (KYN/TRP) ratio in Long COVID (LC) patients versus normal controls.

**Table 3.**
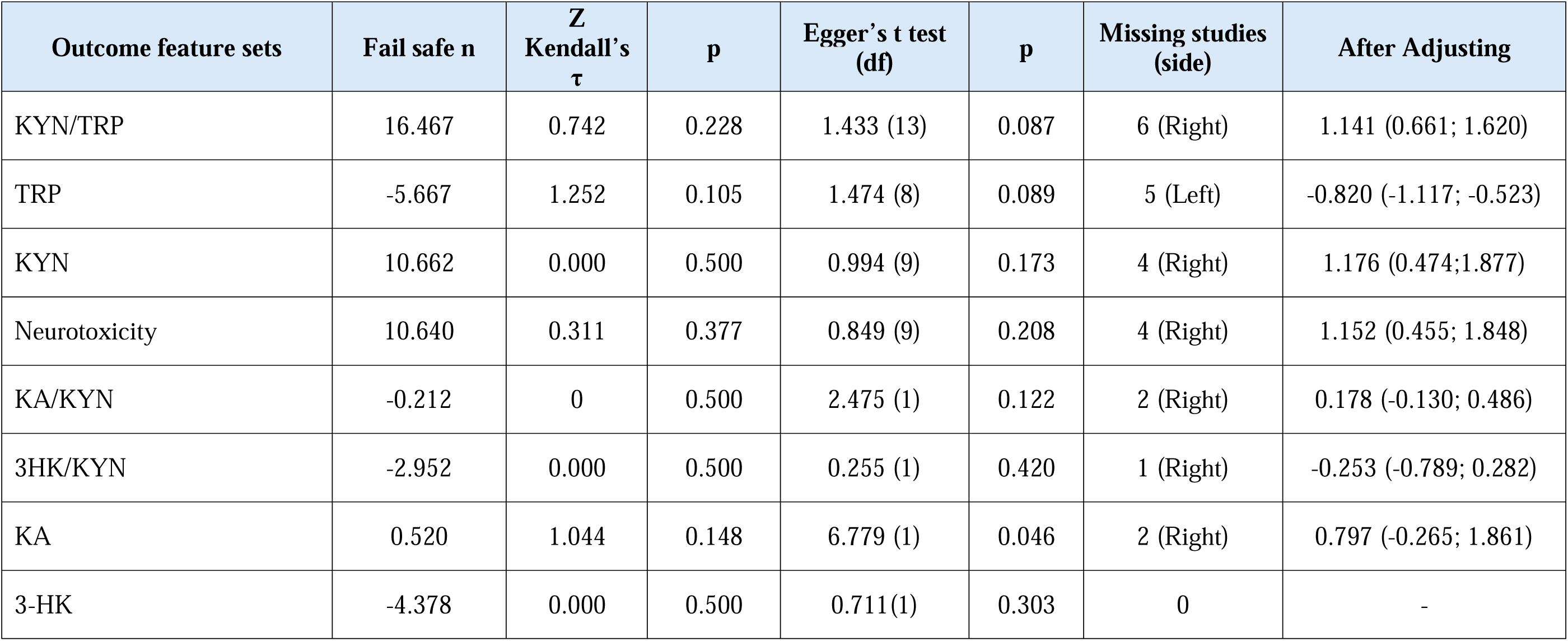
Results on publication bias.

| Outcome feature sets | Fail safe n | Z<br>Kendall's<br>$\tau$ | p | Egger's t test<br>(df) | p | Missing studies<br>(side) | After Adjusting |
| --- | --- | --- | --- | --- | --- | --- | --- |
| KYN/TRP | 16.467 | 0.742 | 0.228 | 1.433 (13) | 0.087 | 6 (Right) | 1.141 (0.661; 1.620) |
| TRP | -5.667 | 1.252 | 0.105 | 1.474 (8) | 0.089 | 5 (Left) | -0.820 (-1.117; -0.523) |
| KYN | 10.662 | 0.000 | 0.500 | 0.994 (9) | 0.173 | 4 (Right) | 1.176 (0.474;1.877) |
| Neurotoxicity | 10.640 | 0.311 | 0.377 | 0.849 (9) | 0.208 | 4 (Right) | 1.152 (0.455; 1.848) |
| KA/KYN | -0.212 | 0 | 0.500 | 2.475 (1) | 0.122 | 2 (Right) | 0.178 (-0.130; 0.486) |
| 3HK/KYN | -2.952 | 0.000 | 0.500 | 0.255 (1) | 0.420 | 1 (Right) | -0.253 (-0.789; 0.282) |
| KA | 0.520 | 1.044 | 0.148 | 6.779 (1) | 0.046 | 2 (Right) | 0.797 (-0.265; 1.861) |
| 3-HK | -4.378 | 0.000 | 0.500 | 0.711(1) | 0.303 | 0 | - |

**Table 4.**
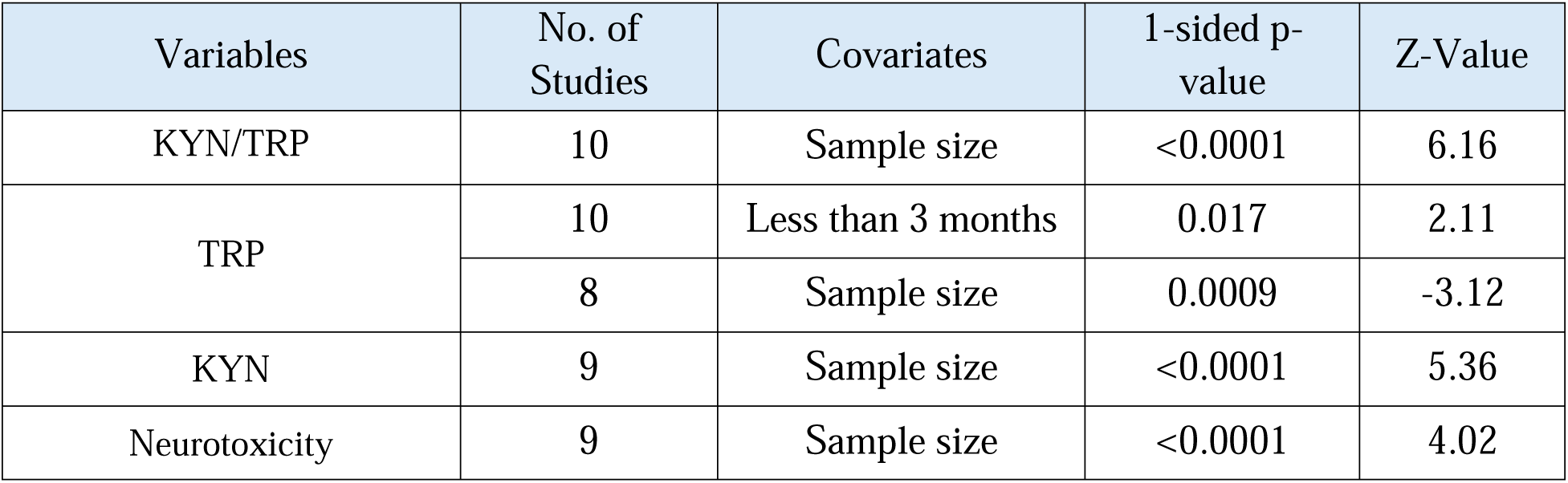
Results of Meta-regression.

#### TRP

Ten studies provided data on TRP levels, showing three studies with CIs entirely below zero and none exclusively above zero (**Table 1**). Seven studies had overlapping CIs, with six indicating an SMD below zero and one above. Our analysis, presented in **Table 2** and **Figure 3**, demonstrated significantly reduced TRP levels in LC patients versus controls. The publication bias analysis revealed 5 studies missing on the left side of the funnel plot (**Table 3**); adjusting for these studies reduced the effect size to -0.820, indicating a further reduction.

**Figure 3:**
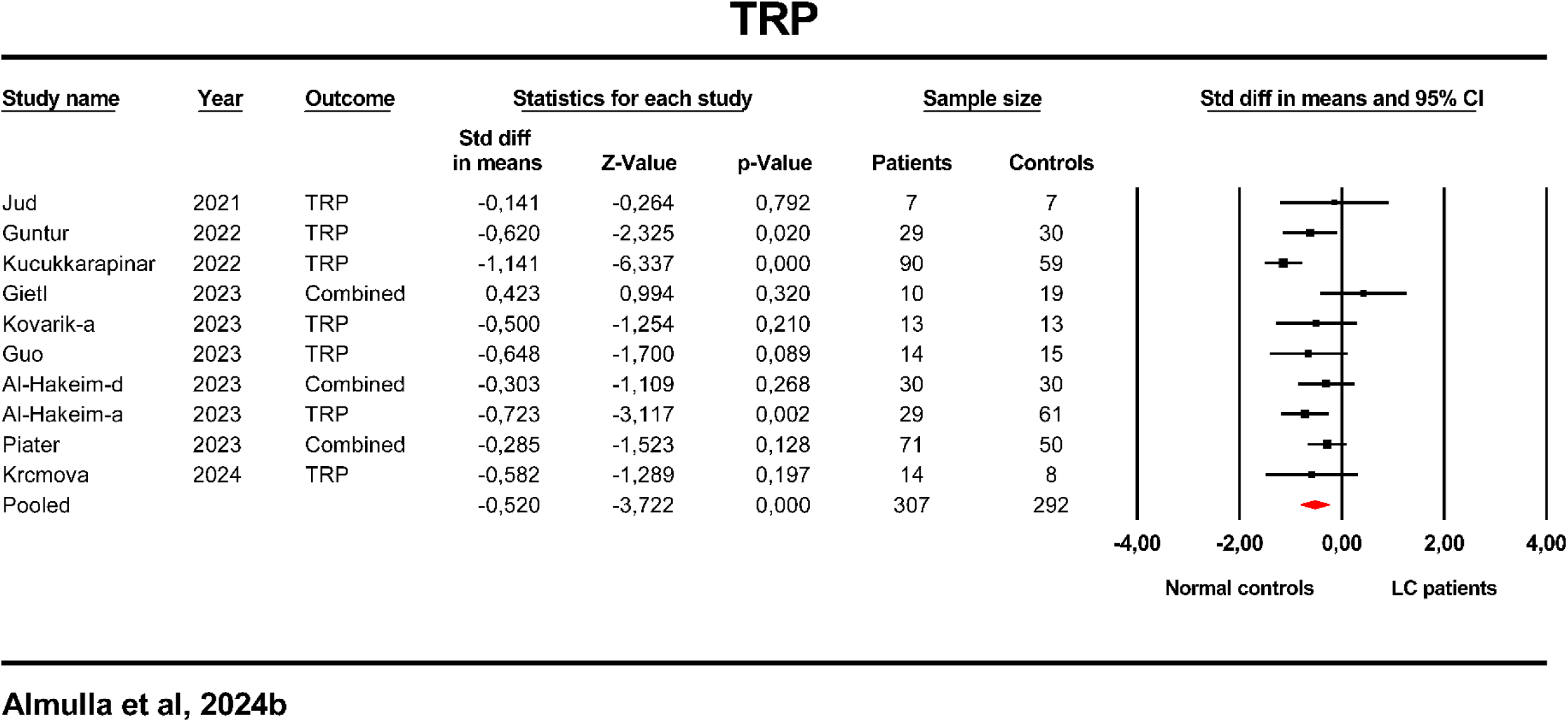
Forest plot with the results of a meta-analysis performed on the tryptophan (TRP) in Long COVID (LC) patients versus normal controls.

#### KYN

Eleven studies assessed KYN levels, with no studies showing CIs solely below zero and five with CIs entirely above (**Table 1**). Six studies had overlapping CIs, with all six indicating a SMD below zero and one documenting a SMD above zero. Our findings, detailed in **Table 2** and **Figure 4**, found no significant change in KYN levels in LC patients. However, publication bias was detected with four studies missing on the right side of the funnel plot (**Table 3**); adjustments resulted in an effect size increase to 1.176, marking a significant elevation.

**Figure 4:**
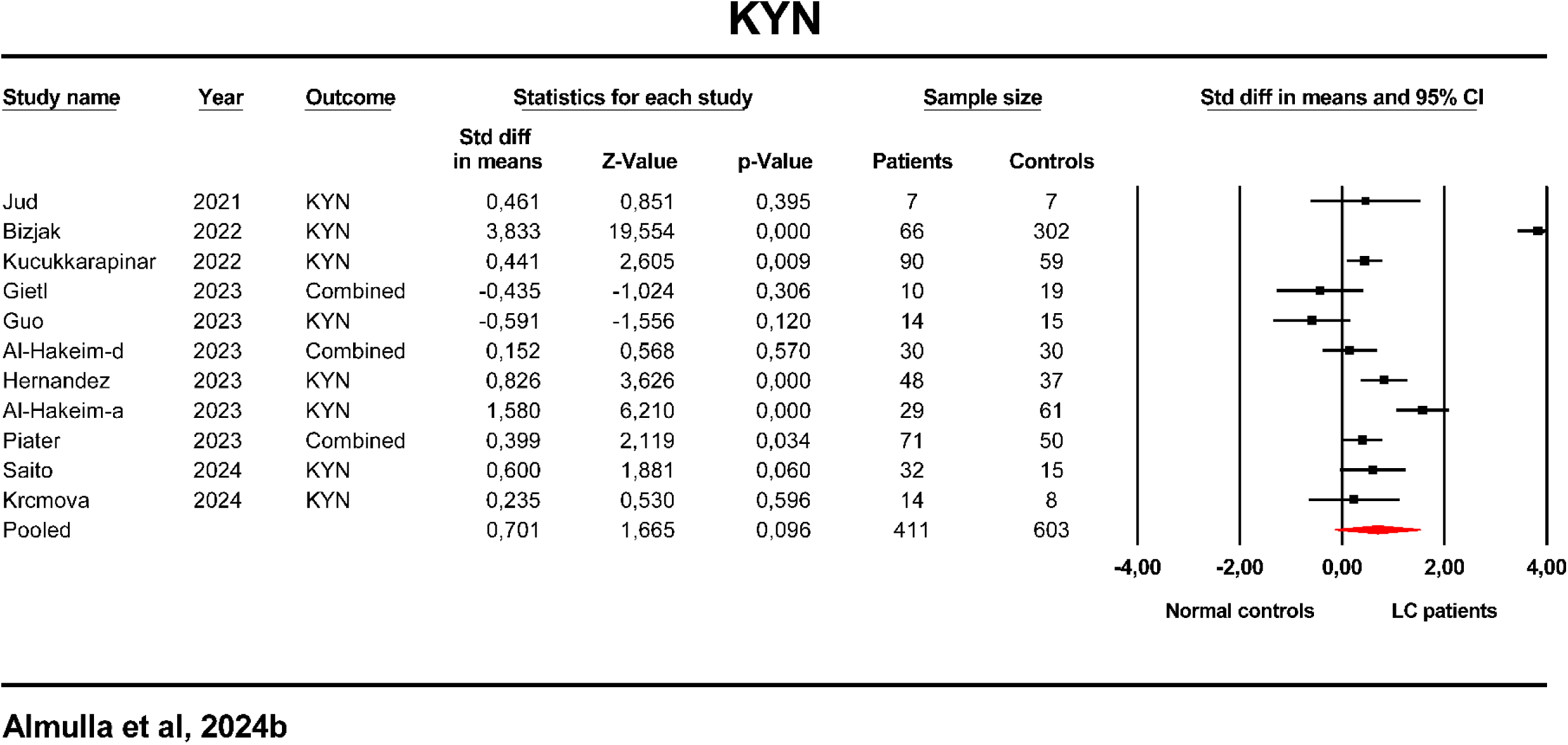
Forest plot with the results of a meta-analysis performed on the kynurenine (KYN) in Long COVID (LC) patients versus normal controls.

#### TRYCAT-related Neurotoxicity

**Table 1** indicated that eleven studies examining neurotoxicity indices showed no CIs completely below zero, but four with CIs above zero. Seven studies had intersecting CIs, with two indicating an SMD below zero and five above. Significant increases in neurotoxicity were observed in LC patients compared to controls, as shown in **Table 2** and **Figure 5**. Publication bias uncovered four studies missing on the right side of the funnel plot; after adjusting for these studies, the effect size increased to 1.118, marking a significant rise.

**Figure 5:**
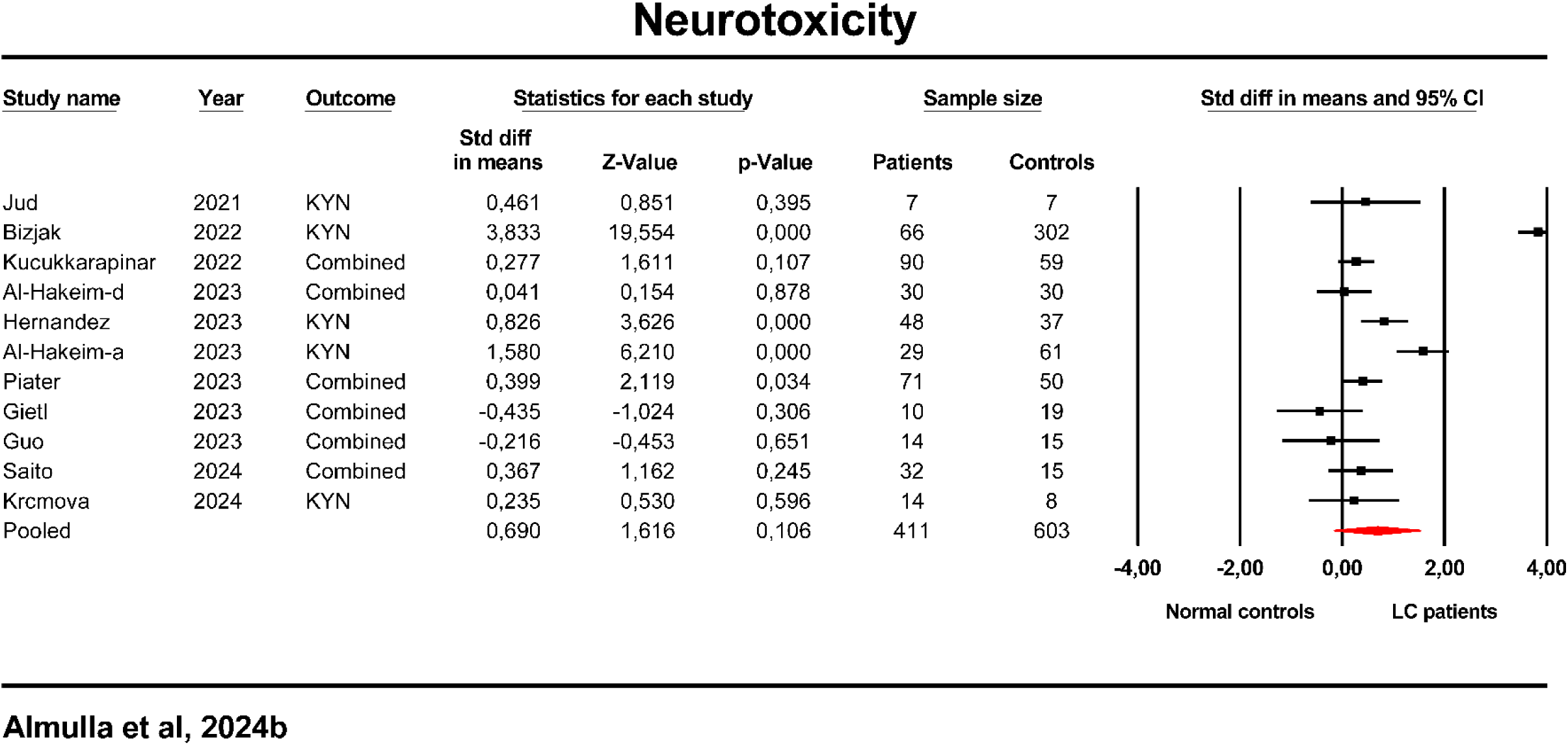
Forest plot with the results of a meta-analysis performed on the neurotoxicity in Long COVID (LC) patients versus normal controls.

#### Secondary Outcome Variables KA/KYN Ratio

This analysis covered three studies on the KA/KYN ratio (**Table 1**). The ratio showed no significant changes in LC patients compared to controls, as detailed in **Table 2** and the ESF, Figure 1. Publication bias analysis detected two missing studies on the right side of the funnel plot, but adjustments did not alter the non-significant results (**Table 3**).

#### 3-HK/KYN Ratio

We examined the 3-HK/KYN ratio across four studies as demonstrated in Table 1. Two of these studies presented CIs entirely below zero, whereas none showed CIs exclusively above zero. Additionally, two studies demonstrated overlapping CIs, with one study reporting a SMD below zero and the other above zero. Notably, a significant reduction in the 3-HK/KYN ratio was observed in patients with LC compared to control subjects, as displayed in Table 2 and ESF Figure 5. Table 3 revealed no evidence of bias.

#### KA

Three studies examining KA levels indicated no significant variation in LC patients (**Table 1**). This was corroborated by findings in **Table 2** and the ESF, Figure 2. Publication bias analysis identifying two missing studies on the right side of the funnel plot. Adjustments for these studies did not change the non-significant outcome (**Table 3**).

#### 3-HK

An assessment of 3-HK levels across four studies revealed no significant changes in LC patients versus controls (Table 1 and ESF, Figure 2). One study was found missing from the left side of the funnel plot (Table 3) but including this study did not significantly alter the effect size, leaving the results statistically insignificant.

### Meta-regression analysis

A meta-regression analysis was conducted to elucidate the specific factors contributing to heterogeneity observed in studies examining TRP and TRYCATs in LC patients. The results indicated that sample sizes had a significant influence on most of the outcomes including TRP, KYN/TRP ratio, KYN and neurotoxicity index, as detailed in ESF, Table 6. In addition, post-COVID period lower than 3 months after acute infection exerts significant impact of TRP results.

## Discussion

### Activation of IDO enzyme in LC disease

The current study’s first finding reveals that LC patients exhibit a heightened KYN/TRP ratio compared to normal controls, suggesting upregulation of IDO, the key enzyme within the TRYCAT pathway. These findings align with our prior research showing increased IDO activity in severe LC (Al-Hakeim, Abed et al. 2023, Al-Hakeim, Khairi Abed et al. 2023) and are corroborated by studies reporting similar elevations in KYN/TRP ratios (Guo, Appelman et al. 2023, Piater, Gietl et al. 2023, Krčmová, Javorská et al. 2024). Additionally, Guo et al. noted elevated IDO2 enzyme expression in white blood cells of LC patients, alongside reduced intracellular TRP levels in peripheral blood mononuclear cells, associated with decreased mitochondrial function and increased autophagy (Guo, Appelman et al. 2023), further implicating the TRYCAT pathway in LC’s pathophysiology.

The activation of the TRYCAT pathway, a primary metabolic route for TRP, results in its depletion and an increase in TRP catabolites in LC patients’ blood. This study found a pronounced decrease in TRP levels and a rise in KYN concentrations among LC patients compared to normal controls, findings that are echoed by a considerable volume of preceding research (Bizjak, Stangl et al. 2022, Kucukkarapinar, Yay-Pence et al. 2022, Guo, Appelman et al. 2023, Piater, Gietl et al. 2023). Notably, our prior meta-analyses on TRP and the TRYCAT pathway in the context of acute COVID-19 identified a significant TRP reduction and TRYCAT pathway activation, particularly pronounced in patients with severe presentations of the disease (Almulla, Supasitthumrong et al. 2022). The current findings suggest ongoing TRYCAT pathway activity, leading to reduced TRP and elevated KYN levels 3-6 months post-recovery, underscoring its potential significance in the pathophysiology of LC.

The reduction in TRP levels and the activation of the TRYCAT pathway in LC are primarily driven by the concomitant activation of immune-inflammatory and oxidative and nitrosative stress (O&NS) pathways. This correlation suggests these stress pathways act as key activators of the rate-limiting enzyme in the TRYCAT pathway, IDO, as previously evidenced (Maes, Leonard et al. 2011, Maes 2015, Al-Hakeim, Al-Rubaye et al. 2022, Almulla, Al-Hakeim et al. 2023). Recent comprehensive meta-analyses have highlighted the activation of the IRS pathway, M1 macrophages, and T helper cells (Th17, Th1, Th2), alongside immune-related neurotoxicity in LC, characterized by elevated levels of relevant cytokines, chemokines, and growth factors (Abbas, Yanin et al. 2024). Furthermore, our investigations have linked LC with activated O&NS pathways, as indicated by increased O&NS biomarkers (malonaldehyde, myeloperoxidase, protein carbonyl) and decreased antioxidant defense components (glutathione peroxidase, zinc) (Al-Hakeim, Al-Rubaye et al. 2022).

Metabolites of the TRYCAT pathway, notably KYN, have been implicated in immune system regulation (Maes et al., 2011). These metabolites possess the capability to attenuate inflammatory responses and modulate the activities of diverse immune cells, offering potential benefits in mitigating excessive inflammation during viral infections (Mándi and Vécsei 2012). Likewise, lowered TRP levels serve as a first-line innate immune defense against infections (Maes et al., 2011). Nevertheless, TRP depletion in airway epithelial-like cells has been associated with increased secretion of IL-6 and CXCL8, a response that can be reversed by TRP supplementation (van Wissen, Snoek et al. 2002).

Recent meta-analytical studies have identified significant decreases in TRP in conditions such as depression and schizophrenia, while no major alterations in TRYCAT levels could be established. This observation suggests that the heightened activity of the TRYCAT pathway is more indicative of acute inflammatory states, exemplified by severe acute COVID-19 infections (Almulla, Supasitthumrong et al. 2022) and interferon (IFN)-induced depression (Maes, Bonaccorso et al. 2001), than of mild chronic inflammatory conditions like major depression (Almulla, Thipakorn et al. 2022a, Almulla, Thipakorn et al. 2022b), schizophrenia (Almulla, Vasupanrajit et al. 2022), autism spectrum disorder (Almulla, Thipakorn et al. 2023), and Alzheimer’s disease (Almulla, Supasitthumrong et al. 2022). Consequently, the observed increase in TRYCAT pathway activity in LC may indicate a continued state of hyperinflammation.

Some authors have posited that the severity of inflammation (as indicated by elevated PBT and hypoxia) during the acute phase of COVID-19 infection could impact the metabolism of TRP in LC, potentially increasing the activity of IDO (Al-Hakeim, Khairi Abed et al. 2023). Bizjak et al. investigated levels of KYN in LC patients, uncovering sustained elevations in both serum and saliva KYN levels (Bizjak, Stangl et al. 2022). These findings indicate that KYN measurements in serum and saliva may serve as indicators of both the immediate and prolonged pathophysiological impacts of SARS-CoV-2 infection, offering potential as valuable biomarkers for the diagnosis and monitoring of the disease (Bizjak, Stangl et al. 2022). Furthermore, a notable decrease in blood albumin—a protein that binds TRP—has been identified as a key factor contributing to TRP depletion in conditions with moderately chronic inflammation, such as depression and schizophrenia. In individuals recovering from acute COVID-19, a significant reduction in serum albumin levels was observed three months post-infection (Gameil, Marzouk et al. 2021), potentially exacerbating TRP deficiency in LC patients.

### KAT and KMO activities in LC

The present investigation’s secondary key discovery is that the KA/KYN and 3-HK/KYN ratios remained unchanged in LC compared to normal individuals. This finding suggests a lack of significant activation in the KAT and KMO enzymes, as these ratios serve as markers for the enzymatic activities of KAT and KMO, respectively (Almulla, Vasupanrajit et al. 2022). Accordingly, our analysis did not reveal notable alterations in the levels of KA and 3-HK, the enzymatic products of KAT and KMO, respectively. This aligns with outcomes from previous research examining levels of KA and 3-HK in LC patients of varying severity (Al-Hakeim, Khairi Abed et al. 2023). Conversely, studies by Guo et al., and Kucukkarapinar et al., have documented a significant reduction in 3-HK levels in individuals post-COVID-19 (Kucukkarapinar, Yay-Pence et al. 2022, Guo, Appelman et al. 2023), while KA levels were found to be significantly elevated in patients who had recovered from COVID-19, assessed six months following the acute phase of infection (Kucukkarapinar, Yay-Pence et al. 2022).

Considering the neuroprotective role of KA, its diminished synthesis in LC could potentially underscore the prominence of neurotoxic TRYCATs. Previous investigations have identified analogous patterns regarding KAT enzyme activity in patients with acute COVID-19 (Almulla, Supasitthumrong et al. 2022). Notably, KA concentrations were found to be significantly elevated in the serum of individuals with acute COVID-19, whereas plasma levels exhibited no significant changes (Almulla, Supasitthumrong et al. 2022), highlighting the variability of KA levels across different biological mediums.

### TRYCAT-related neurotoxicity in LC disease

The present study’s third key finding indicates no significant association between LC disease and TRYCATs-related neurotoxicity, as determined by an index derived from the cumulative neurotoxic TRYCATs, including KYN, 3-HK, 3-HAA, QA, XA, and PA. However, individual analyses reported elevated KYN levels without significant changes in 3-HK levels among LC patients, and comprehensive assessments of other neurotoxic TRYCATs such as 3-HAA, QA, XA, and PA was not feasible due to the scarcity of studies. Previous research has identified specific TRYCATs, particularly 3-HK and QA, as contributors to neuro-oxidative stress, facilitating oxidative damage to cellular structures and promoting lipid peroxidation (Smith, Smith et al. 2009, Reyes Ocampo, Lugo Huitrón et al. 2014, Almulla and Maes 2022). The increased production of neurotoxic TRYCATs has been implicated in the development of neuropsychiatric symptoms observed in conditions such as major depression, bipolar disorder, schizophrenia, anxiety, somatization disorder, and chronic fatigue syndrome, as well as in neurocognitive impairments (Maes, Galecki et al. 2011, Cordeiro, Guimaraes et al. 2014, Ormstad, Simonsen et al. 2020, Milaneschi, Allers et al. 2021).

Research has highlighted that TRYCATs with neurotoxic effects, notably KYN and 3-HK, are implicated in the development of major depression induced by IFN-α (Maes, Bonaccorso et al. 2001, Bonaccorso, Marino et al. 2002). The emergence of depressive symptoms following IFN-α treatment is more closely linked to the synthesis of KYN and an elevated KYN/KA ratio than to decreased TRP levels (Maes, Bonaccorso et al. 2001, Bonaccorso, Marino et al. 2002, Wichers, Koek et al. 2005, Maes 2015). Existing literature indicates that QA and 3-HK are implicated in neurodegenerative processes and may contribute to the cognitive impairments and brain structural changes observed in bipolar disorder (Birner, Platzer et al. 2017). Furthermore, increased production of KYN has been associated with effects that are depressogenic, anxiogenic, excitotoxic, and neurotoxic (Wichers, Koek et al. 2005, Maes 2011), potentially impacting the physio-affective phenome of LC (Almulla, Al-Hakeim et al. 2023).

Our meta-regression analysis identified sample size as a significant factor affecting TRP and TRYCAT pathway findings, underscoring the importance of utilizing well-powered case-control designs to explore TRP and TRYCAT dynamics in LC.

## Limitations

This study’s conclusions are drawn within the context of certain limitations. Primarily, the analysis did not extend to solitary levels of downstream TRP metabolites such as AA, 3-HAA, QA, XA, and PA due to a scarcity of pertinent literature. Additionally, variations between serum/plasma and the central nervous system (CNS) in TRP and TRYCATs, as previously documented (Almulla, Vasupanrajit et al. 2022), were not evaluated in the current study owing to the limited number of serum/plasma studies and the absence of CNS research. Furthermore, the potential effects of medications prescribed for LC symptoms were not explored, as no studies provided relevant information. Lastly, the impact of vaccination status and the type of vaccine administered could not be assessed due to unavailable data.

## Conclusion

This meta-analysis suggests that the pathophysiology of LC disease is characterized by a sustained reduction in TRP levels, activation of the TRYCAT pathway, and increased KYN levels 3-6 months after full recovery from acute COVID infection. Strategies aimed at replenishing diminished TRP levels and normalizing the activity of the TRYCAT pathway may offer viable therapeutic avenues for managing LC.

## Data Availability

Upon request, and subject to all authors' full consent, the corresponding author, MM, will respond to valid requests for the dataset employed in this meta-analysis, which will be provided in Excel file format.

## Ethical approval and consent to participate

Not applicable.

## Consent for publication

Not applicable.

## Author’s contributions

AA and MM were responsible for designing the present study, while YT and AA collected the data. YT and AA independently verified the data accuracy. AA and MM conducted the statistical analysis. All authors have actively participated in the drafting and revising of the manuscript and have provided their approval for the submission of the final version.

## Acknowledgments

Not applicable.

## References

Abbas, F. A., T. Yanin, Z. Bo, V. Aristo and M. Michael (2024). “Immune activation and immune-associated neurotoxicity in Long-COVID: A systematic review and meta-analysis of 82 studies comprising 58 cytokines/chemokines/growth factors.” medRxiv: 2024.2002.2008.24302516.

Al-Hadrawi, D. S., H. T. Al-Rubaye, A. F. Almulla, H. K. Al-Hakeim and M. Maes (2022). “Lowered oxygen saturation and increased body temperature in acute COVID-19 largely predict chronic fatigue syndrome and affective symptoms due to Long COVID: A precision nomothetic approach.” Acta Neuropsychiatrica: 1–12.

Al-Hakeim, H. K., A. K. Abed, A. F. Almulla, S. Rouf Moustafa and M. Maes (2023). “Anxiety due to Long COVID is partially driven by activation of the tryptophan catabolite (TRYCAT) pathway.” Asian Journal of Psychiatry 88: 103723.

Al-Hakeim, H. K., H. T. Al-Rubaye, D. S. Al-Hadrawi, A. F. Almulla and M. Maes (2022). “Long-COVID post-viral chronic fatigue and affective symptoms are associated with oxidative damage, lowered antioxidant defenses and inflammation: a proof of concept and mechanism study.” Molecular Psychiatry.

Al-Hakeim, H. K., H. T. Al-Rubaye, A. F. Almulla, D. S. Al-Hadrawi and M. Maes (2023) “Chronic Fatigue, Depression and Anxiety Symptoms in Long COVID Are Strongly Predicted by Neuroimmune and Neuro-Oxidative Pathways Which Are Caused by the Inflammation during Acute Infection.” Journal of Clinical Medicine 12 DOI: 10.3390/jcm12020511.

Al-Hakeim, H. K., H. T. Al-Rubaye, A. S. Jubran, A. F. Almulla, S. R. Moustafa and M. Maes (2023). “Increased insulin resistance due to Long COVID is associated with depressive symptoms and partly predicted by the inflammatory response during acute infection.” Braz J Psychiatry.

Al-Hakeim, H. K., A. Khairi Abed, S. Rouf Moustafa, A. F. Almulla and M. Maes (2023). “Tryptophan catabolites, inflammation, and insulin resistance as determinants of chronic fatigue syndrome and affective symptoms in long COVID.” Frontiers in Molecular Neuroscience 16.

Al-Jassas, H. K., H. K. Al-Hakeim and M. Maes (2022). “Intersections between pneumonia, lowered oxygen saturation percentage and immune activation mediate depression, anxiety, and chronic fatigue syndrome-like symptoms due to COVID-19: A nomothetic network approach.” J Affect Disord 297: 233–245.

Almulla, A. F., H. K. Al-Hakeim and M. Maes (2023). “Chronic fatigue and affective symptoms in acute and long COVID are attributable to immune-inflammatory pathways.” Psychiatry Clin Neurosci 77(2): 125–126.

Almulla, A. F. and M. Maes (2023). “Although serotonin is not a major player in depression, its precursor is.” Molecular Psychiatry 28(8): 3155–3156.

Almulla, A. F., M. Maes, Z. Bo, K. A.-H. Hussein and V. Aristo (2023). “Brain-targeted autoimmunity is strongly associated with Long COVID and its chronic fatigue syndrome as well as its affective symptoms.” medRxiv: 2023.2010.2004.23296554.

Almulla, A. F., T. Supasitthumrong, A. Amrapala, C. Tunvirachaisakul, A. K. A. Jaleel, G. Oxenkrug, H. K. Al-Hakeim and M. Maes (2022). “The Tryptophan Catabolite or Kynurenine Pathway in Alzheimer’s Disease: A Systematic Review and Meta-Analysis.” J Alzheimers Dis 88(4): 1325–1339.

Almulla, A. F., T. Supasitthumrong, C. Tunvirachaisakul, A. A. A. Algon, H. K. Al-Hakeim and M. Maes (2022). “The tryptophan catabolite or kynurenine pathway in COVID-19 and critical COVID-19: a systematic review and meta-analysis.” BMC Infectious Diseases 22(1): 615.

Almulla, A. F., Y. Thipakorn, C. Tunvirachaisakul and M. Maes (2023). “The tryptophan catabolite or kynurenine pathway in autism spectrum disorder; a systematic review and meta-analysis.” Autism Res 16(12): 2302–2315.

Almulla, A. F., Y. Thipakorn, A. Vasupanrajit, A. A. Abo Algon, C. Tunvirachaisakul, A. A. Hashim Aljanabi, G. Oxenkrug, H. K. Al-Hakeim and M. Maes (2022a). “The tryptophan catabolite or kynurenine pathway in major depressive and bipolar disorder: A systematic review and meta-analysis.” Brain, Behavior, & Immunity - Health 26: 100537.

Almulla, A. F., Y. Thipakorn, A. Vasupanrajit, C. Tunvirachaisakul, G. Oxenkrug, H. K. Al-Hakeim and M. Maes (2022b). “The Tryptophan Catabolite or Kynurenine Pathway in a Major Depressive Episode with Melancholia, Psychotic Features and Suicidal Behaviors: A Systematic Review and Meta-Analysis.” Cells 11(19).

Almulla, A. F., A. Vasupanrajit, C. Tunvirachaisakul, H. K. Al-Hakeim, M. Solmi, R. Verkerk and M. Maes (2022). “The tryptophan catabolite or kynurenine pathway in schizophrenia: meta-analysis reveals dissociations between central, serum, and plasma compartments.” Molecular Psychiatry.

Almulla, F. A. and M. Maes (2022). “The Tryptophan Catabolite or Kynurenine Pathway’s Role in Major Depression.” Current Topics in Medicinal Chemistry 22(21): 1731–1735.

Andres-Rodriguez, L., X. Borras, A. Feliu-Soler, A. Perez-Aranda, N. Angarita-Osorio, P. Moreno-Peral, J. Montero-Marin, J. Garcia-Campayo, A. F. Carvalho and M. Maes (2020). “Peripheral immune aberrations in fibromyalgia: A systematic review, meta-analysis and meta-regression.” Brain, behavior, and immunity 87: 881–889.

Badenoch, J. B., E. R. Rengasamy, C. Watson, K. Jansen, S. Chakraborty, R. D. Sundaram, D. Hafeez, E. Burchill, A. Saini, L. Thomas, B. Cross, C. K. Hunt, I. Conti, S. Ralovska, Z. Hussain, M. Butler, T. A. Pollak, I. Koychev, B. D. Michael, H. Holling, T. R. Nicholson, J. P. Rogers and A. G. Rooney (2022). “Persistent neuropsychiatric symptoms after COVID-19: a systematic review and meta-analysis.” Brain Commun 4(1): fcab297.

Birner, A., M. Platzer, S. A. Bengesser, N. Dalkner, F. T. Fellendorf, R. Queissner, R. Pilz, P. Rauch, A. Maget, C. Hamm, S. Herzog-Eberhard, H. Mangge, D. Fuchs, N. Moll, S. Zelzer, G. Schütze, M. Schwarz, B. Reininghaus, H. P. Kapfhammer and E. Z. Reininghaus (2017). “Increased breakdown of kynurenine towards its neurotoxic branch in bipolar disorder.” PLoS One 12(2): e0172699.

Bizjak, D. A., M. Stangl, N. Börner, F. Bösch, J. Durner, G. Drunin, J.-L. Buhl and D. Abendroth (2022). “Kynurenine serves as useful biomarker in acute, Long- and Post-COVID-19 diagnostics.” Frontiers in Immunology 13.

Bonaccorso, S., V. Marino, M. Biondi, F. Grimaldi, F. Ippoliti and M. Maes (2002). “Depression induced by treatment with interferon-alpha in patients affected by hepatitis C virus.” Journal of affective disorders 72(3): 237–241.

Ceban, F., S. Ling, L. M. W. Lui, Y. Lee, H. Gill, K. M. Teopiz, N. B. Rodrigues, M. Subramaniapillai, J. D. Di Vincenzo, B. Cao, K. Lin, R. B. Mansur, R. C. Ho, J. D. Rosenblat, K. W. Miskowiak, M. Vinberg, V. Maletic and R. S. McIntyre (2022). “Fatigue and cognitive impairment in Post-COVID-19 Syndrome: A systematic review and meta-analysis.” Brain Behav Immun 101: 93–135.

Cohen, J. (2013). Statistical power analysis for the behavioral sciences, Routledge.

Cordeiro, L., J. Guimaraes, S. Wanner, R. La Guardia, R. Miranda, U. Marubayashi and D. Soares (2014). “Inhibition of tryptophan hydroxylase abolishes fatigue induced by central tryptophan in exercising rats.” Scandinavian Journal of Medicine & Science in Sports 24(1): 80–88.

Duval, S. and R. Tweedie (2000a). “Trim and fill: A simple funnel-plot-based method of testing and adjusting for publication bias in meta-analysis.” Biometrics 56(2): 455–463.

Duval, S. and R. Tweedie (2000b). “A Nonparametric “Trim and Fill” Method of Accounting for Publication Bias in Meta-Analysis.” Journal of the American Statistical Association 95(449): 89–98.

Gameil, M. A., R. E. Marzouk, A. H. Elsebaie and S. E. Rozaik (2021). “Long-term clinical and biochemical residue after COVID-19 recovery.” Egyptian Liver Journal 11(1): 74.

Gietl, M., F. Burkert, S. Seiwald, A. Böhm, S. Hofer, J. M. Gostner, T. Piater, S. Geisler, G. Weiss, J. Loeffler-Ragg, T. Sonnweber, I. Tancevski, A. Pizzini, S. Sahanic, D. Fuchs, R. Bellmann-Weiler and K. Kurz (2023). “Interferon-gamma Mediated Metabolic Pathways in Hospitalized Patients During Acute and Reconvalescent COVID-19.” Int J Tryptophan Res 16: 11786469231154244.

Guntur, V. P., T. Nemkov, E. de Boer, M. P. Mohning, D. Baraghoshi, F. I. Cendali, I. San-Millán, I. Petrache and A. D’Alessandro (2022). “Signatures of Mitochondrial Dysfunction and Impaired Fatty Acid Metabolism in Plasma of Patients with Post-Acute Sequelae of COVID-19 (PASC).” Metabolites 12(11).

Guo, L., B. Appelman, K. Mooij-Kalverda, R. H. Houtkooper, M. van Weeghel, F. M. Vaz, A. Dijkhuis, T. Dekker, B. S. Smids, J. W. Duitman, M. Bugiani, P. Brinkman, J. J. Sikkens, H. A. A. Lavell, R. C. I. Wüst, M. van Vugt, R. Lutter, M. A. van Agtmael, A. G. Algera, B. Appelman, F. E. H. P. van Baarle, M. Beudel, H. J. Bogaard, M. Bomers, P. I. Bonta, L. D. J. Bos, M. Botta, J. de Brabander, G. J. de Bree, S. de Bruin, M. Bugiani, E. B. Bulle, O. Chouchane, A. P. M. Cloherty, D. Buis, M. C. F. J. de Rotte, M. Dijkstra, D. A. Dongelmans, R. W. G. Dujardin, P. E. Elbers, L. M. Fleuren, S. E. Geerlings, T. B. H. Geijtenbeek, A. R. J. Girbes, A. Goorhuis, M. P. Grobusch, L. A. Hagens, J. Hamann, V. C. Harris, R. Hemke, S. M. Hermans, L. M. A. Heunks, M. W. Hollmann, J. Horn, J. W. Hovius, M. D. de Jong, R. Koing, E. H. T. Lim, N. van Mourik, J. F. Nellen, E. J. Nossent, F. Paulus, E. Peters, D. Piña-Fuentes, T. van der Poll, B. Preckel, J. M. Prins, S. J. Raasveld, T. D. Y. Reijnders, M. Schinkel, F. A. P. Schrauwen, M. J. Schultz, A. R. Schuurman, J. Schuurmans, K. Sigaloff, M. A. Slim, P. Smeele, M. R. Smit, C. Stijnis, W. Stilma, C. E. Teunissen, P. Thoral, A. M. Tsonas, P. R. Tuinman, M. van der Valk, D. P. Veelo, C. Volleman, H. de Vries, L. A. van Vught, M. van Vugt, D. Wouters, A. H. Zwinderman, M. C. Brouwer, W. J. Wiersinga, A. P. J. Vlaar and D. van de Beek (2023). “Prolonged indoleamine 2,3-dioxygenase-2 activity and associated cellular stress in post-acute sequelae of SARS-CoV-2 infection.” eBioMedicine 94.

Havell, E. A. and J. Vilcek (1972). “Production of high-titered interferon in cultures of human diploid cells.” Antimicrob Agents Chemother 2(6): 476–484.

Higgins, J. P., S. G. Thompson, J. J. Deeks and D. G. Altman (2003). “Measuring inconsistency in meta-analyses.” Bmj 327(7414): 557–560.

Higgins JPT, T. J., Chandler J, Cumpston M, Li T, Page MJ, Welch VA (2019). Cochrane Handbook for Systematic Reviews of Interventions. Chichester (UK), John Wiley & Sons.

Holmes, E., J. Wist, R. Masuda, S. Lodge, P. Nitschke, T. Kimhofer, R. L. Loo, S. Begum, B. Boughton, R. Yang, A. C. Morillon, S. T. Chin, D. Hall, M. Ryan, S. H. Bong, M. Gay, D. W. Edgar, J. C. Lindon, T. Richards, B. B. Yeap, S. Pettersson, M. Spraul, H. Schaefer, N. G. Lawler, N. Gray, L. Whiley and J. K. Nicholson (2021). “Incomplete Systemic Recovery and Metabolic Phenoreversion in Post-Acute-Phase Nonhospitalized COVID-19 Patients: Implications for Assessment of Post-Acute COVID-19 Syndrome.” J Proteome Res 20(6): 3315–3329.

Jud, P., P. Gressenberger, V. Muster, A. Avian, A. Meinitzer, H. Strohmaier, H. Sourij, R. B. Raggam, M. H. Stradner, U. Demel, H. H. Kessler, K. Eller and M. Brodmann (2021). “Evaluation of Endothelial Dysfunction and Inflammatory Vasculopathy After SARS-CoV-2 Infection—A Cross-Sectional Study.” Frontiers in Cardiovascular Medicine 8.

Kovarik, J. J., A. Bileck, G. Hagn, S. M. Meier-Menches, T. Frey, A. Kaempf, M. Hollenstein, T. Shoumariyeh, L. Skos, B. Reiter, M. C. Gerner, A. Spannbauer, E. Hasimbegovic, D. Schmidl, G. Garhöfer, M. Gyöngyösi, K. G. Schmetterer and C. Gerner (2023). “A multi-omics based anti-inflammatory immune signature characterizes long COVID-19 syndrome.” iScience 26(1): 105717.

Krčmová, L. K., L. Javorská, K. Matoušová, P. Šmahel, M. Skála, M. Kopecký, C. Suwanvecho, N. Přívratská, D. Turoňová and B. Melichar (2024). “Evaluation of inflammatory biomarkers and vitamins in hospitalized patients with SARS-CoV-2 infection and post-COVID syndrome.” Clin Chem Lab Med.

Kucukkarapinar, M., A. Yay-Pence, Y. Yildiz, M. Buyukkoruk, G. Yaz-Aydin, T. S. Deveci-Bulut, O. Gulbahar, E. Senol and S. Candansayar (2022). “Psychological outcomes of COVID-19 survivors at sixth months after diagnose: the role of kynurenine pathway metabolites in depression, anxiety, and stress.” J Neural Transm (Vienna) 129(8): 1077–1089.

López-Hernández, Y., J. Monárrez-Espino, D. A. G. López, J. Zheng, J. C. Borrego, C. Torres-Calzada, J. P. Elizalde-Díaz, R. Mandal, M. Berjanskii, E. Martínez-Martínez, J. A. López and D. S. Wishart (2023). “The plasma metabolome of long COVID patients two years after infection.” Scientific Reports 13(1): 12420.

Lutter, R., M. van Wissen, T. Roger, P. Bresser, K. van der Sluijs, M. Nijhuis and H. M. Jansen (2003). “Mechanisms that potentially underlie virus-induced exaggerated inflammatory responses by airway epithelial cells.” Chest 123(3 Suppl): 391s-392s.

Maes, M. (2011). “Depression is an inflammatory disease, but cell-mediated immune activation is the key component of depression.” Progress in Neuro-Psychopharmacology and Biological Psychiatry 35(3): 664–675.

Maes, M. (2015). “A review on citation amnesia in depression and inflammation research.” Neuro Endocrinol Lett 36(1): 1–6.

Maes, M., H. T. Al-Rubaye, A. F. Almulla, D. S. Al-Hadrawi, K. Stoyanova, M. Kubera and H. K. Al-Hakeim (2022) “Lowered Quality of Life in Long COVID Is Predicted by Affective Symptoms, Chronic Fatigue Syndrome, Inflammation and Neuroimmunotoxic Pathways.” International Journal of Environmental Research and Public Health 19 DOI: 10.3390/ijerph191610362.

Maes, M., S. Bonaccorso, V. Marino, A. Puzella, M. Pasquini, M. Biondi, M. Artini, C. Almerighi and H. Meltzer (2001). “Treatment with interferon-alpha (IFNα) of hepatitis C patients induces lower serum dipeptidyl peptidase IV activity, which is related to IFNα-induced depressive and anxiety symptoms and immune activation.” Molecular psychiatry 6(4): 475–480.

Maes, M., P. Galecki, R. Verkerk and W. Rief (2011). “Somatization, but not depression, is characterized by disorders in the tryptophan catabolite (TRYCAT) pathway, indicating increased indoleamine 2, 3-dioxygenase and lowered kynurenine aminotransferase activity.” Neuroendocrinol Lett 32(3): 264–273.

Maes, M., B. E. Leonard, A. M. Myint, M. Kubera and R. Verkerk (2011). “The new ’5-HT’ hypothesis of depression: cell-mediated immune activation induces indoleamine 2,3-dioxygenase, which leads to lower plasma tryptophan and an increased synthesis of detrimental tryptophan catabolites (TRYCATs), both of which contribute to the onset of depression.” Prog Neuropsychopharmacol Biol Psychiatry 35(3): 702–721.

Mándi, Y. and L. Vécsei (2012). “The kynurenine system and immunoregulation.” J Neural Transm (Vienna) 119(2): 197–209.

Merlo, L. M. F., J. B. DuHadaway, J. D. Montgomery, W. D. Peng, P. J. Murray, G. C. Prendergast, A. J. Caton, A. J. Muller and L. Mandik-Nayak (2020). “Differential Roles of IDO1 and IDO2 in T and B Cell Inflammatory Immune Responses.” Front Immunol 11: 1861.

Merlo, L. M. F., E. Pigott, J. B. DuHadaway, S. Grabler, R. Metz, G. C. Prendergast and L. Mandik-Nayak (2014). “IDO2 is a critical mediator of autoantibody production and inflammatory pathogenesis in a mouse model of autoimmune arthritis.” J Immunol 192(5): 2082–2090.

Milaneschi, Y., K. A. Allers, A. T. Beekman, E. J. Giltay, S. Keller, R. A. Schoevers, S. D. Süssmuth, H. G. Niessen and B. W. Penninx (2021). “The association between plasma tryptophan catabolites and depression: The role of symptom profiles and inflammation.” Brain, behavior, and immunity 97: 167–175.

Nalbandian, A., K. Sehgal, A. Gupta, M. V. Madhavan, C. McGroder, J. S. Stevens, J. R. Cook, A. S. Nordvig, D. Shalev, T. S. Sehrawat, N. Ahluwalia, B. Bikdeli, D. Dietz, C. Der-Nigoghossian, N. Liyanage-Don, G. F. Rosner, E. J. Bernstein, S. Mohan, A. A. Beckley, D. S. Seres, T. K. Choueiri, N. Uriel, J. C. Ausiello, D. Accili, D. E. Freedberg, M. Baldwin, A. Schwartz, D. Brodie, C. K. Garcia, M. S. V. Elkind, J. M. Connors, J. P. Bilezikian, D. W. Landry and E. Y. Wan (2021). “Post-acute COVID-19 syndrome.” Nat Med 27(4): 601–615.

NICE, N. I. f. H. a. C. E. (2020, 11 Nov. 2021). “COVID-19 rapid guideline: managing the long-term effects of COVID-19.” Retrieved 27 March, 2022, from https://www.nice.org.uk/guidance/ng188.

Ormstad, H., C. S. Simonsen, L. Broch, M. Maes, G. Anderson and E. G. Celius (2020). “Chronic fatigue and depression due to multiple sclerosis: Immune-inflammatory pathways, tryptophan catabolites and the gut-brain axis as possible shared pathways.” Multiple sclerosis and related disorders 46: 102533.

Page, M. J., J. E. McKenzie, P. M. Bossuyt, I. Boutron, T. C. Hoffmann, C. D. Mulrow, L. Shamseer, J. M. Tetzlaff, E. A. Akl, S. E. Brennan, R. Chou, J. Glanville, J. M. Grimshaw, A. Hróbjartsson, M. M. Lalu, T. Li, E. W. Loder, E. Mayo-Wilson, S. McDonald, L. A. McGuinness, L. A. Stewart, J. Thomas, A. C. Tricco, V. A. Welch, P. Whiting and D. Moher (2021). “The PRISMA 2020 statement: An updated guideline for reporting systematic reviews.” PLoS medicine 18(3): e1003583–e1003583.

Phillips, S. and M. A. Williams (2021). “Confronting Our Next National Health Disaster - Long-Haul Covid.” N Engl J Med 385(7): 577–579.

Piater, T., M. Gietl, S. Hofer, J. M. Gostner, S. Sahanic, I. Tancevski, T. Sonnweber, A. Pizzini, A. Egger, H. Schennach, J. Loeffler-Ragg, G. Weiss and K. Kurz (2023). “Persistent Symptoms and IFN-γ-Mediated Pathways after COVID-19.” J Pers Med 13(7).

Reyes Ocampo, J., R. Lugo Huitrón, D. González-Esquivel, P. Ugalde-Muñiz, A. Jiménez-Anguiano, B. Pineda, J. Pedraza-Chaverri, C. Ríos and V. Pérez de la Cruz (2014). “Kynurenines with neuroactive and redox properties: relevance to aging and brain diseases.” Oxid Med Cell Longev 2014: 646909.

Roger, T., T. Out, N. Mukaida, K. Matsushima, H. Jansen and R. Lutter (1998). “Enhanced AP-1 and NF-kappaB activities and stability of interleukin 8 (IL-8) transcripts are implicated in IL-8 mRNA superinduction in lung epithelial H292 cells.” Biochem J 330 (**Pt 1**)(Pt 1): 429–435.

Saito, S., S. Shahbaz, X. Luo, M. Osman, D. Redmond, J. W. Cohen Tervaert, L. Li and S. Elahi (2024). “Metabolomic and immune alterations in long COVID patients with chronic fatigue syndrome.” Front Immunol 15: 1341843.

Smith, A. J., R. A. Smith and T. W. Stone (2009). “5-Hydroxyanthranilic Acid, a Tryptophan Metabolite, Generates Oxidative Stress and Neuronal Death via p38 Activation in Cultured Cerebellar Granule Neurones.” Neurotoxicity Research 15(4): 303–310.

van Wissen, M., M. Snoek, B. Smids, H. M. Jansen and R. Lutter (2002). “IFN-gamma amplifies IL-6 and IL-8 responses by airway epithelial-like cells via indoleamine 2,3-dioxygenase.” J Immunol 169(12): 7039–7044.

Wichers, M. C., G. Koek, G. Robaeys, R. Verkerk, S. Scharpe and M. Maes (2005). “IDO and interferon-α-induced depressive symptoms: a shift in hypothesis from tryptophan depletion to neurotoxicity.” Molecular psychiatry 10(6): 538–544.

World Health Organization, W. (2021). “A clinical case definition of post COVID-19 condition by a Delphi consensus 6 October.” WHO.

