## supplementary file for "The tryptophan catabolite or kynurenine pathway in Long COVID disease: A systematic review and meta-analysis"

SHORT TITLE: Kynurenine pathway in Long COVID disease

1. Medical Laboratory Technology Department, College of Medical Technology, The Islamic University, Najaf, Iraq.

2. Department of Psychiatry, Faculty of Medicine, Chulalongkorn University, Bangkok, Thailand.

3. Sichuan Provincial Center for Mental Health, Sichuan Provincial People’s Hospital, School of Medicine, University of Electronic Science and Technology of China, Chengdu 610072, China

4. Key Laboratory of Psychosomatic Medicine, Chinese Academy of Medical Sciences, Chengdu, 610072, China

5. Immunosciences Lab, Inc., Los Angeles, CA 90035, USA.

6. Cyrex Laboratories, LLC, Phoenix, AZ 85034, USA.

7. Department of Psychiatry, Medical University of Plovdiv, Plovdiv, Bulgaria.

8. Research Institute, Medical University of Plovdiv, Plovdiv, Bulgaria.

9. Strategic Research and Innovation Program for the Development of MU - PLOVDIV–(SRIPD-MUP), European Union – NextGenerationEU.

10. Cognitive Impairment and Dementia Research Unit, Faculty of Medicine, Chulalongkorn University, Bangkok, Thailand.

11. Cognitive Fitness and Biopsychological Technology Research Unit, Faculty of Medicine. Chulalongkorn University, Bangkok, 10330, Thailand, Bangkok, 10330, Thailand.

12. Kyung Hee University, 26 Kyungheedae-ro, Dongdaemun-gu, Seoul 02447, Korea.

* **Corresponding author:**

Prof. Dr. Michael Maes, M.D., Ph.D.

Sichuan Provincial Center for Mental Health

Sichuan Provincial People’s Hospital,

School of Medicine,

University of Electronic Science and Technology of China

Chengdu 610072

China

e-mail addresses:

<https://scholar.google.co.th/citations?user=1wzMZ7UAAAAJ&hl=th&oi=ao>

Highly cited author: 2003-2023 (ISI, Clarivate)

ScholarGPS: Worldwide #1 in molecular neuroscience; #1/4 in pathophysiology

Expert worldwide medical expertise ranking, Expertscape (December 2022), worldwide:

#1 in CFS, #1 in oxidative stress, #1 in encephalomyelitis, #1 in nitrosative stress, #1 in nitrosation, #1 in tryptophan, #1 in aromatic amino acids, #1 in stress (physiological), #1 in neuroimmune; #2 in bacterial translocation; #3 in inflammation, #4-5: in depression, fatigue and psychiatry.

**ESF. Table 1.** Search sentences and terms were used in each database.

| **Database Name** | **Search Sentence** | **No. of Articles** |
| --- | --- | --- |
| **PubMed/Medline** | ("Long COVID" OR "Post COVID-19 Syndrome" OR "Post-acute Sequelae of SARS-CoV-2 infection" OR "COVID-19 long-term effects") AND ("Kynurenine Pathway" OR "Tryptophan Metabolism" OR "Kynurenine" OR "Tryptophan" OR "Kynurenic Acid" OR "Quinolinic Acid" OR "Picolinic Acid" OR "3-Hydroxykynurenine" OR "Anthranilic Acid" OR "3-Hydroxyanthranilic Acid" OR "Xanthurenic Acid" OR "Tryptophan Catabolites") | **20** |
|  | ("Long COVID" OR "Post COVID-19 Syndrome" OR "PASC" OR "Post-acute Sequelae of SARS-CoV-2 infection") AND ("Kynurenine Pathway" OR "Tryptophan Metabolism") AND ("Kynurenine" OR "Tryptophan" OR "Kynurenic Acid" OR "Quinolinic Acid" OR "Picolinic Acid" OR "3-Hydroxykynurenine" OR "Anthranilic Acid" OR "3-Hydroxyanthranilic Acid" OR "Xanthurenic Acid") | **6** |
|  | (COVID-19[MeSH Terms] OR "Long COVID" OR "Post COVID Syndrome" OR "Post-acute Sequelae of SARS-CoV-2 infection") AND (Tryptophan[MeSH Terms] OR Kynurenine[MeSH Terms] OR "Kynurenine Pathway" OR "Tryptophan Metabolism" OR "Kynurenic Acid" OR "Quinolinic Acid" OR "Picolinic Acid" OR "3-Hydroxykynurenine" OR "Anthranilic Acid" OR "3-Hydroxyanthranilic Acid" OR "Xanthurenic Acid") | **83** |
|  | ("COVID-19"[Mesh] OR "SARS-CoV-2"[Mesh] OR "Long COVID" OR "Post-acute Sequelae of SARS-CoV-2 infection" OR "Post COVID-19 condition") AND ("Tryptophan Oxygenase"[Mesh] OR "Tryptophan"[Mesh] OR "Kynurenine"[Mesh] OR "Kynurenine Pathway" OR "Tryptophan Catabolites" OR "Kynurenic Acid" OR "Quinolinic Acid" OR "Picolinic Acid" OR "3-Hydroxykynurenine" OR "Anthranilic Acid" OR "3-Hydroxyanthranilic Acid" OR "Xanthurenic Acid") | **79** |
| **Google Scholar** | ("Long COVID" OR "Post COVID" OR "Post-acute sequelae of SARS-CoV-2 infection" OR "PASC") AND ("kynurenine pathway" OR "tryptophan metabolism" OR "tryptophan catabolites" OR "kynurenic acid" OR "quinolinic acid" OR "picolinic acid" OR "3-hydroxykynurenine" OR "anthranilic acid" OR "kynurenine" OR "3-hydroxyanthranilic acid" OR "xanthurenic acid") | **1110** |
|  | ("Long COVID" OR "Post-COVID-19 syndrome" OR "PASC" OR "COVID-19 sequelae") AND ("kynurenine pathway" OR "tryptophan metabolism" OR "tryptophan degradation" OR "KYNA" OR "QUIN" OR "PIC" OR "3-HK" OR "AA" OR "3-HAA" OR "XA") | **7220** |
| **SciFinder** | "Long COVID" OR "Post COVID-19 Syndrome" OR "Post-acute Sequelae of SARS-CoV-2 Infection" OR "COVID-19 long-term effects") AND ("Kynurenine Pathway" OR "Tryptophan Metabolism" OR "Kynurenine" OR "Tryptophan" OR "Kynurenic Acid" OR "Quinolinic Acid" OR "Picolinic Acid" OR "3-Hydroxykynurenine" OR "Anthranilic Acid" OR "3-Hydroxyanthranilic Acid" OR "Xanthurenic Acid" OR "Tryptophan Catabolites" | **36** |

**ESF. Table 2.** Immune cofounder’s scale (ICS) applied from Andrés-Rodríguez. et al.. 2019

| **Methodological quality of the study** | |
| --- | --- |
| **1** | Study sample ≥ 128 participants including patients and controls (1= Yes. 0 = No) |
| **2** | Did the study control the results for potential confounders (e.g.. age. BMI. gender. race)? (1= Yes. 0 = No) |
| **3** | Were participants with Long COVID and controls age- and-gender-matched or was there a statistical control? (1= Yes. 0 = No) |
| **4** | Was the time of sample collection specified (e.g.. morning vs. evening)? (1= Yes. 0 = No) |
| **5** | Were participants with Long COVID free of immunomodulatory drugs including anti-cytokines. glucocorticoids. immunoglobulins. and immunosuppressants. or was there a medication washout period. or was drug intake statistically controlled for? (1= Yes. 0 = No) |
| **6** | Were participants with Long COVID free of antidepressants and mood stabilizers or were the data statistically controlled for? (1= Yes. 0 = No) |
| **7** | Reporting either the manufacturer of the test or detection limit and coefficients of variation (1= Yes. 0 = No) |
| **8** | Reporting how data under detection limit were handled (1 = Yes. 0 = No) |
| **9** | Reporting % of the sample under detection limit (1=Yes. 0= No) |
| **10** | Reporting blood fraction (serum. plasma. culture supernatant or whole blood) (1= Yes. 0 = No) |
| **Total quality score (10 points)** | |
| **Biomarker confounders red points**  *The red points should not be given if the item is statistically controlled for* | |
| **1** | 3 red points for comorbid illnesses such as autoimmune disorders & other immune disorders including rheumatoid arthritis. psoriasis. inflammatory bowel disease. chronic obstructive pulmonary disease. multiple sclerosis |
| **2** | 3 red points for use of recreational drugs such as methamphetamine or opioids |
| **3** | 2 red points when groups were not matched for age |
| **4** | 2 red points when groups were not matched for sex |
| **5** | 2 red points for medication use as for example immunomodulators |
| **6** | 2 red points for early traumatic life events |
| **7** | 2 red points for shift work and primary sleep disorders |
| **8** | 1.5 red points for use of antipsychotics |
| **9** | 1 red point for more common systemic immune disorders including diabetes type 1/2. essential hypertension. metabolic syndrome |
| **10** | 1 red point for not fasting (8 hours before blood extraction) |
| **11** | 1 red point for use of omega-3 and antioxidant supplements |
| **12** | 1 red point when data were not controlled for body mass index |
| **13** | 1 red point when data were not controlled for physical activity or sedentary life |
| **14** | 1 red point when data were not controlled for smoking |
| **15** | 1 red point for use of oral contraceptives or NSAIDs |
| **16** | 0.5 red points when data were not controlled for ethnicity in countries such as US. Brazil |
| **17** | 0.5 red points when data were not controlled for seasonality |
| **18** | 0.5 red points when data were not controlled for diurnal variation (8-10 a.m. versus all other time points) |
|  | **Total red point score (26 points)** |

**ESF. Table 3.** PRISMA checklist

| **Section/topic** | **#** | **Checklist item** | **Reported on page #** |
| --- | --- | --- | --- |
| **TITLE** | | | |
| Title | 1 | Identify the report as a systematic review. meta-analysis. or both. | 1 |
| **ABSTRACT** | | | |
| Structured summary | 2 | Provide a structured summary including. as applicable: background; objectives; data sources; study eligibility criteria. participants. and interventions; study appraisal and synthesis methods; results; limitations; conclusions and implications of key findings; systematic review registration number. | 3 |
| **INTRODUCTION** | | | |
| Rationale | 3 | Describe the rationale for the review in the context of what is already known. | 5 |
| Objectives | 4 | Provide an explicit statement of questions being addressed with reference to participants. interventions. comparisons. outcomes. and study design (PICOS). | 8 |
| **METHODS** | | | |
| Protocol and registration | 5 | Indicate if a review protocol exists. if and where it can be accessed (e.g.. Web address). and. if available. provide registration information including registration number. | 9 |
| Eligibility criteria | 6 | Specify study characteristics (e.g.. PICOS. length of follow-up) and report characteristics (e.g.. years considered. language. publication status) used as criteria for eligibility. giving rationale. | 10 |
| Information sources | 7 | Describe all information sources (e.g.. databases with dates of coverage. contact with study authors to identify additional studies) in the search and date last searched. | 9 |
| Search | 8 | Present full electronic search strategy for at least one database. including any limits used. such that it could be repeated. | ESF. Table 1. page 3 |
| Study selection | 9 | State the process for selecting studies (i.e.. screening. eligibility. included in systematic review. and. if applicable. included in the meta-analysis). | 10 |
| Data collection process | 10 | Describe method of data extraction from reports (e.g.. piloted forms. independently. in duplicate) and any processes for obtaining and confirming data from investigators. | 11 |
| Data items | 11 | List and define all variables for which data were sought (e.g.. PICOS. funding sources) and any assumptions and simplifications made. | 11 |
| Risk of bias in individual studies | 12 | Describe methods used for assessing risk of bias of individual studies (including specification of whether this was done at the study or outcome level). and how this information is to be used in any data synthesis. | 12 |
| Summary measures | 13 | State the principal summary measures (e.g.. risk ratio. difference in means). | 12 |
| Synthesis of results | 14 | Describe the methods of handling data and combining results of studies. if done. including measures of consistency (e.g.. I^2^) for each meta-analysis. | 12 |
| Risk of bias across studies | 15 | Specify any assessment of risk of bias that may affect the cumulative evidence (e.g.. publication bias. selective reporting within studies). | Table 3. page 71 |
| Additional analyses | 16 | Describe methods of additional analyses (e.g.. sensitivity or subgroup analyses. meta-regression). if done. indicating which were pre-specified. | 12 |
| **RESULTS** | | |  |
| Study selection | 17 | Give numbers of studies screened. assessed for eligibility. and included in the review. with reasons for exclusions at each stage. ideally with a flow diagram. | 14 |
| Study characteristics | 18 | For each study. present characteristics for which data were extracted (e.g.. study size. PICOS. follow-up period) and provide the citations. | ESF. Table 2 page |
| Risk of bias within studies | 19 | Present data on risk of bias of each study and. if available. any outcome level assessment (see item 12). | Table 3. page 71 |
| Results of individual studies | 20 | For all outcomes considered (benefits or harms). present. for each study: (a) simple summary data for each intervention group (b) effect estimates and confidence intervals. ideally with a forest plot. | Table 1. page 66 |
| Synthesis of results | 21 | Present results of each meta-analysis done. including confidence intervals and measures of consistency. | Table 2. page 68 |
| Risk of bias across studies | 22 | Present results of any assessment of risk of bias across studies (see Item 15). | Table 3 page 71 |
| Additional analysis | 23 | Give results of additional analyses. if done (e.g.. sensitivity or subgroup analyses. meta-regression [see Item 16]). | Table 2. page 68 |
| **DISCUSSION** | | |  |
| Summary of evidence | 24 | Summarize the main findings including the strength of evidence for each main outcome; consider their relevance to key groups (e.g.. healthcare providers. users. and policy makers). | Page 22-31 |
| Limitations | 25 | Discuss limitations at study and outcome level (e.g.. risk of bias). and at review-level (e.g.. incomplete retrieval of identified research. reporting bias). | Page 31-32 |
| Conclusions | 26 | Provide a general interpretation of the results in the context of other evidence. and implications for future research. | Page 32 |
| **FUNDING** | | |  |
| Funding | 27 | Describe sources of funding for the systematic review and other support (e.g.. supply of data); role of funders for the systematic review. | Page 33 |

**ESF. table 4.** Characteristics of the studies included in the systematic reviews and meta-analysis.

| **NO** | **Authors. years** | **Setting** | **Type of case** | **Type of Control** | **Sample Size** | | | **Age** | | **Assessed**  **Biomarkers** | **Specimen** | **Method** | **Quality score** | **Red point score** | **Findnigs** |
| --- | --- | --- | --- | --- | --- | --- | --- | --- | --- | --- | --- | --- | --- | --- | --- |
|  |  |  |  |  | **Cases**  **M/F** | **Control**  **M/F** | **Total**  **M/F** | **Case-Mean (SD)** | **Control- Mean(SD)** |  |  |  |  |  |  |
| 1 | Holmes. Wist et al. 2021 | Australia | Post COVID | Healthy Control | - | - | - | - | - | KYN/TRP | Plasma | LC-MS | 4 | 14,5 | KYN/TRP# |
| 2 | Jud. Gressenberger et al. 2021 | Austria | Post COVID | Controls | 14 7/7 | 14 7/7 | 28 14/14 | 68.7 (12.0) | 30.7 (4.2) | KYN, TRP  ,KYN/TRP | Serum | HPLC | 6 | 9 | KYN#, TRP*  ,KYN/TRP# |
| 3 | Bizjak. Stangl et al. 2022 | Germany | Long COVID | Normal Controls | - | - | - | 66.6 (17.6) | 48.3 (18.3) | KYN | Serum | Spectrophotometer | 5,5 | 10,5 | KYN# |
| 4 | Guntur. Nemkov et al. 2022 | USA | Post COVID | Controls | 29 12/17 | 30 19/11 | 59 31/28 | 42 (13) | 48 (11) | TRP | Plasma | UHPLC-MS | 5 | 10 | TRP* |
| 5 | Kucukkarapinar. Yay-Pence et al. 2022 | Turkey | Post COVID | Controls | 90 39/51 | 59 17/42 | 149 56/93 | 44.5 (15.3) | 30.5 (7.4) | TRP, KYN  KYN/TRP  KA, 3-HK  QA, KA/3-HK,KA/QA | Serum | LC–MS/MS | 5,5 | 12 | TRP*, KYN#  KYN/TRP#  KA#, 3-HK*  QA#, KA/3-HK#,KA/QA# |
| 6 | Al-Hakeim. Abed et al. 2023 | Iraq | Post COVID | Healthy Control | 29 -/- | 61 -/- | 90  -/- | - | - | TRP, KYN ,KYN/TRP | Serum | ELISA | 7 | 7,5 | TRP*, KYN# ,KYN/TRP# |
| 7 | Al-Hakeim. Khairi Abed et al. 2023 | Iraq | Long COVID (2groups) | Healthy Control | 20 12/8 | 30 16/14 | 70 34/36 | 32.5 (8.4) | 29.2 (8.1) | TRP, 3-HK, QA, KA, KYN, KYN/TRP | Serum | ELISA | 7 | 9,5 | TRP, 3-HK#, QA*, KA*, KYN#, KYN/TRP# |
| 8 | Gietl. Burkert et al. 2023 | Austria | Long COVID (3groups) | No symptoms | - | - | - | - | - | KYN, TRP  KYN/TRP | Serum | HPLC | 4 | 14,5 | KYN#, TRP*  KYN/TRP* |
| 9 | Guo. Appelman et al. 2023 | Netherlands | Post COVID | Fully Recovered | 15 3/12 | 14 8/6 | 29 11/18 | 49  (42.5-54) | 31.5  (29-44.25) | TRP, KYN  KA,3-HK  XA, AA, 3-HAA, QA | Plasma | UPLC-MS/MS | 4,5 | 10,5 | TRP*, KYN*  KA*,3-HK*  XA#, AA, 3-HAA#, QA |
| 10 | Kovarik. Bileck et al. 2023 | Austria | Long COVID | Healthy Control | 13 4/9 | 13 6/7 | 26 10/16 | 33  (21-53) | 30  (25-43) | TRP | Plasma | LC-MS/MS | 4,5 | 10,5 | TRP* |
| 11 | López-Hernández. Monárrez-Espino et al. 2023 | Mexico | Post COVID | Controls | 48 28/20 | 37 17/20 | 85 45/40 | 51.5  (43.5–60.8) | 40.5  (37–53.3) | KYN, KYN/TRP | Plasma | LC–MS/MS | 5 | 10 | KYN#, KYN/TRP# |
| 12 | Piater. Gietl et al. 2023 | Austria | Post COVID (2groups) | Published data of Healthy donors | 79 79/0 | 58 58/0 | 137 137/0 | 57.3 (14.1) | 51.1 (11.0) | TRP, KYN, KYN/TRP | Plasma | HPLC | 6 | 13 | TRP*, KYN#, KYN/TRP# |
| 13 | Krčmová. Javorská et al. 2024 | Czech Republic | Post COVID | No Post COVID | 14 7/7 | 8 5/3 | 22 12/10 | 65.5 | 64 | KYN, TRP  KYN/TRP | Serum | HPLC-FLD/PDA | 5 | 12 | KYN#, TRP*  KYN/TRP# |
| 14 | Saito. Shahbaz et al. 2024 | Canada | Long COVID | Healthy Control | 32 8/24 | 15 5/10 | 47 13/34 | 50.5 (12.1) | 47.4 (13.9) | KYN | Plasma | LC-MS | 4,5 | 10,5 | KYN# |

*: Indicates that patients have reduced level of the measured metabolite compared to healthy control

^#:^ Indicates that patients have increased level of the measured metabolites compared to healthy control

TRP: Tryptophan. KYN: Kynurenine. KA: Kynurenic acid. 3HK: 3-Hydroxykynurenine. AA: Anthranilic acid. 3HA: 3-Hydroxyanthranilic acid. XA: Xanthurenic acid. QA: Quinolinic acid. PA: Picolinic acid. HPLC: High performance liquid chromatography. HPLC-MS/MS: High performance liquid chromatography with tandem mass spectrometry. LC-MS/MS: Liquid chromatography with tandem mass spectrometry. UPLC-MS/MS: Ultra performance liquid chromatography with tandem mass spectrometry. LC: Liquid chromatography. HPLC-UV: High perfomance liquid chromatography- Ultra-violate

**ESF. Table 5**. Results of Meta-regression

| Variables | No. of Studies | Covariates | 1-sided p-value | Z-Value |
| --- | --- | --- | --- | --- |
| KYN/TRP | 10 | Sample size | <0.0001 | 6.16 |
| TRP | 10 | Less than 3 months | 0.017 | 2.11 |
|  | 8 | Sample size | 0.0009 | -3.12 |
| KYN | 9 | Sample size | <0.0001 | 5.36 |
| Neurotoxicity | 9 | Sample size | <0.0001 | 4.02 |

TRP: Tryptophan. KYN: Kynurenine.

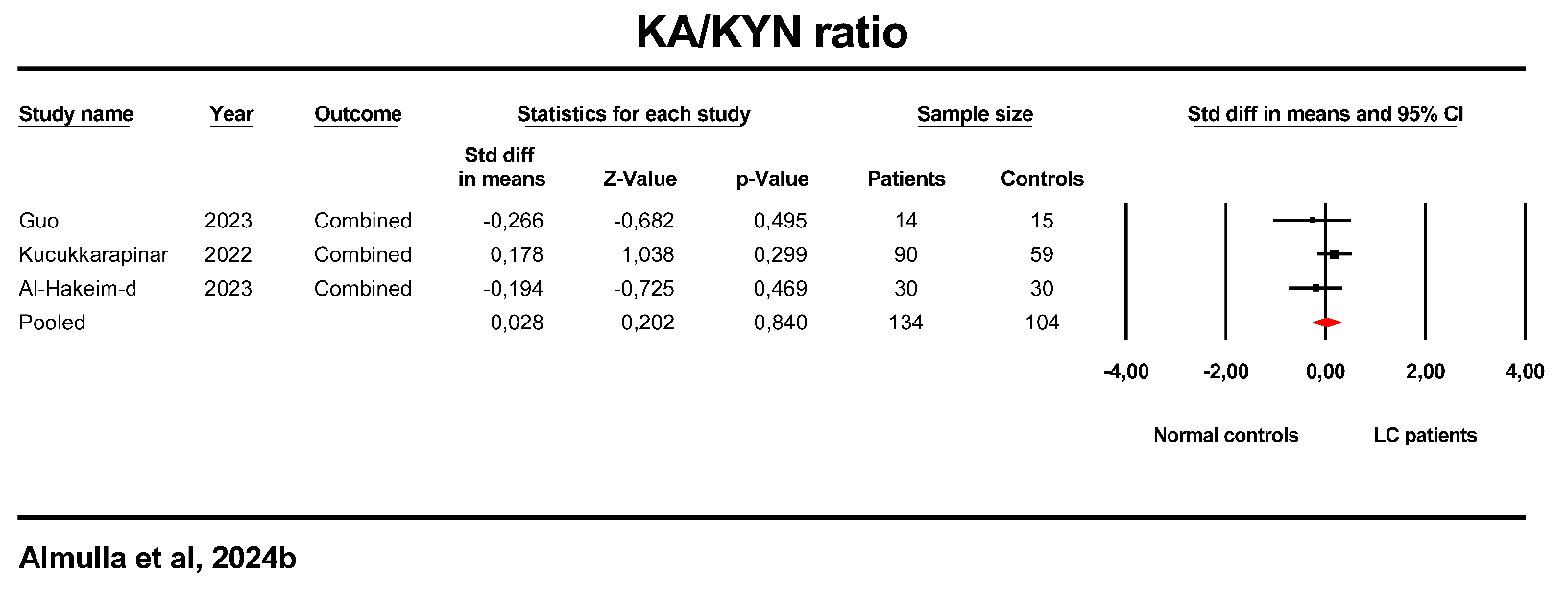

**ESF. Figure 1**. Forest plot of kynurenic acid (KA)/kynurenine (KYN) in the patients with Long COVID (LC) versus healthy control.

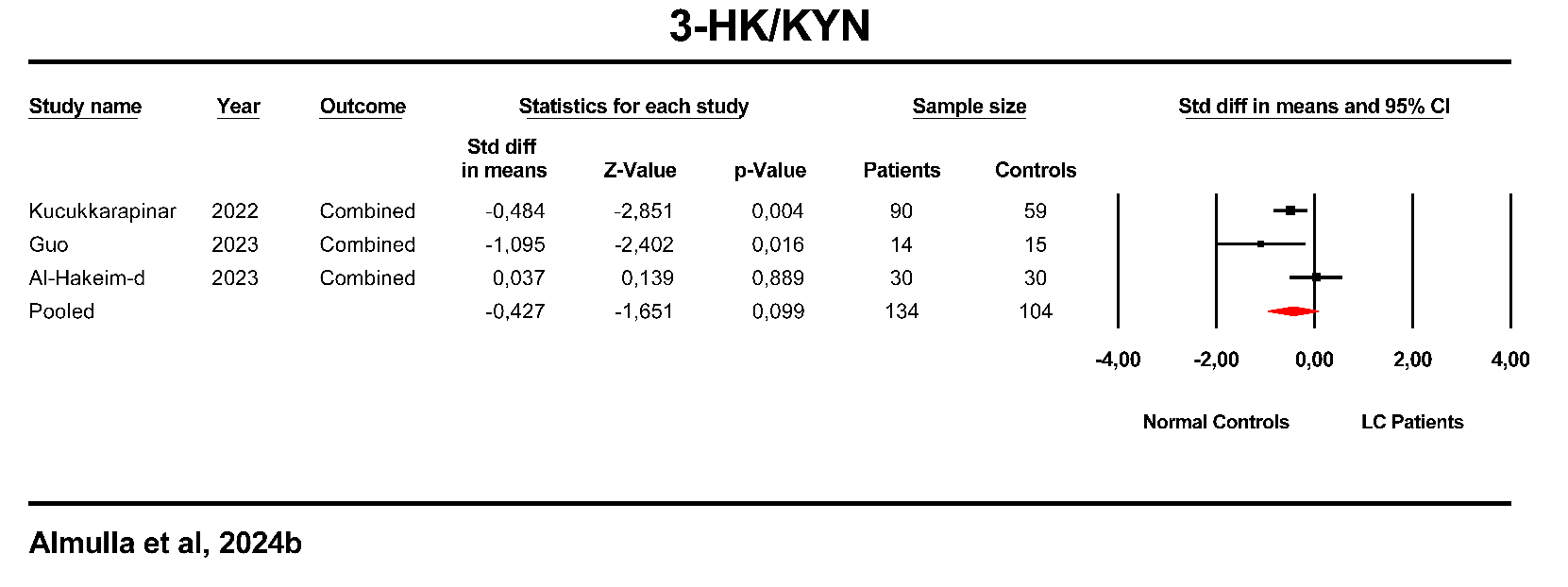

**ESF. Figure 2**. Forest plot of 3-Hydroxykynurenine (3-HK)/kynurenine (KYN) in the patients with Long COVID (LC) versus healthy control.

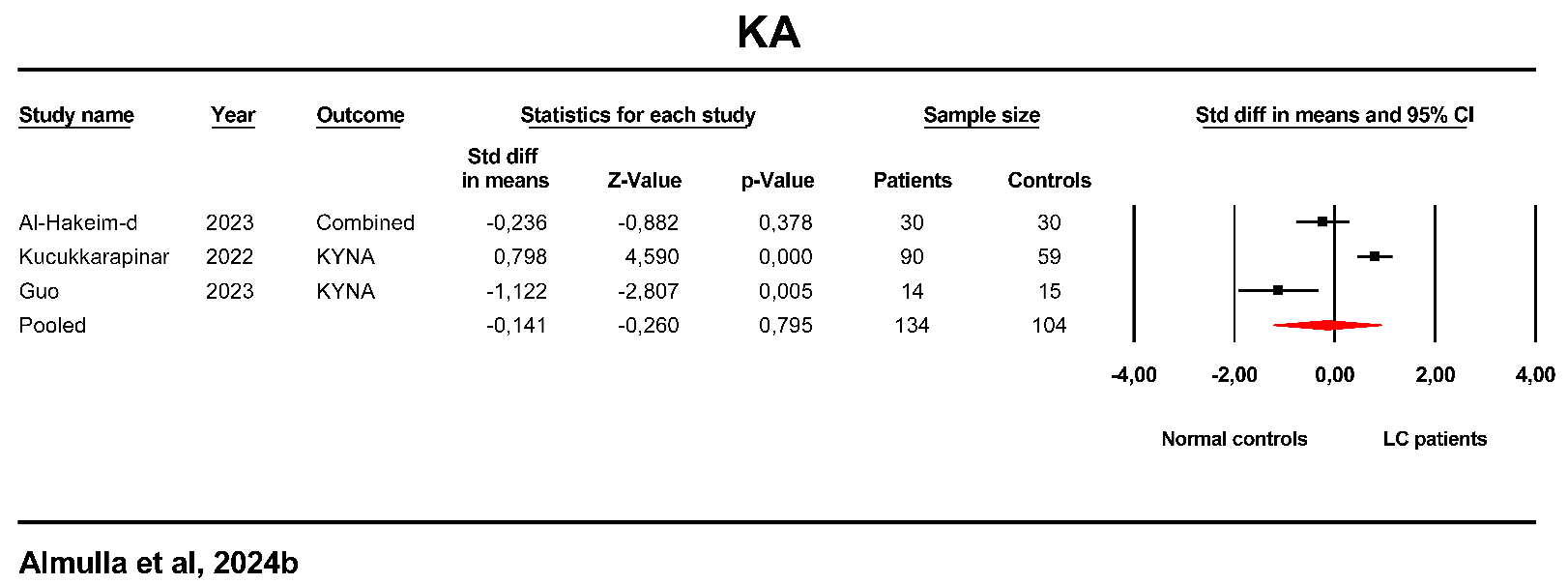

**ESF. Figure 3**. Forest plot of kynurenic acid in the patients with Long COVID (LC) versus healthy control.

.

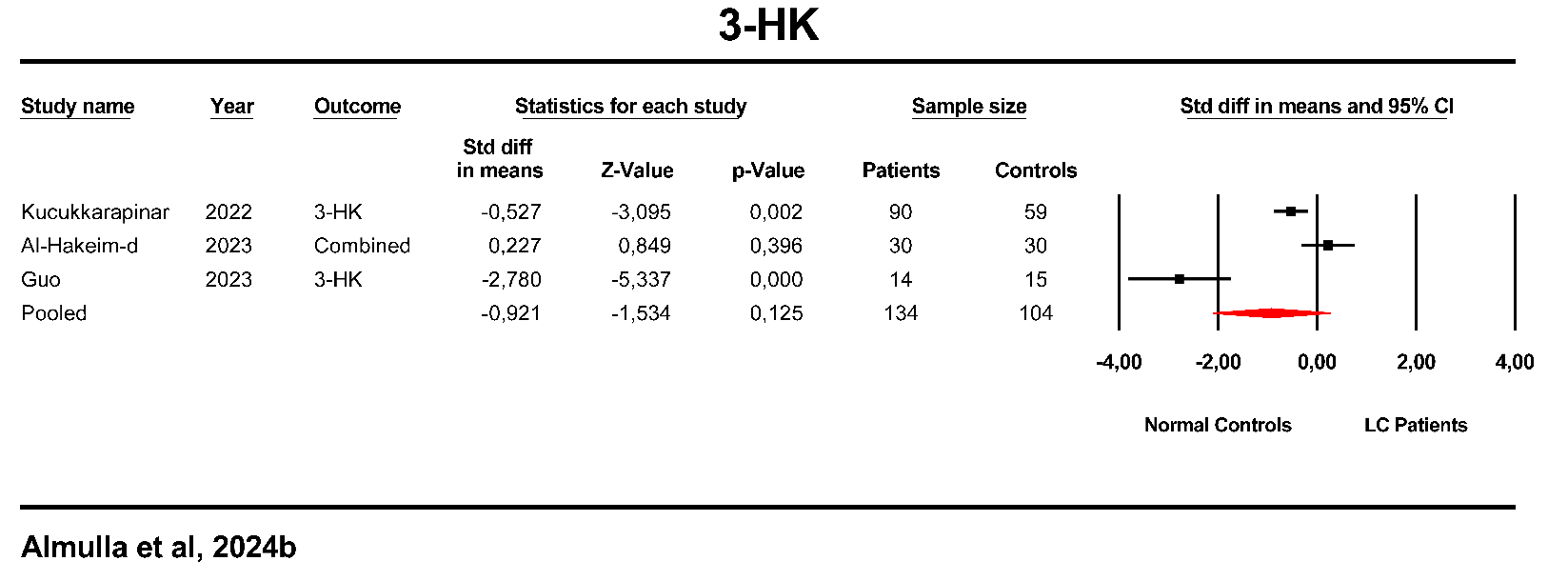

**ESF. Figure 4**.Forest plot of 3-Hydroxykynurenine (3-HK) in the patients with Long COVID (LC) versus healthy control..

**Abbreviation:**

TRP: Tryptophan

KYN: Kynurenine

KA: Kynurenic acid

3HK: 3-Hydroxykynurenine

AA: Anthranilic acid

3HAA: 3-Hydroxyanthranilic acid

XA: Xanthurenic acid

QA: Quinolinic acid

PA: Picolinic acid

IDO: Indoleamine 2.3 dioxygenase

KAT: Kynurenine aminotransferase

KMO: Kynurenine 3-monooxygenase

TRYCATs: Tryptophan Catabolites

TRYCAT pathway: Tryptophan catabolite pathway

SMD: Standardized mean difference

IL-6: Interleukin-6

O&NS: Oxidative and nitrosative stress

MOOSE: Meta-Analyses of Observational Studies in Epidemiology

CSF: Cerebrospinal fluid

SD: Standard definition

IOR: Interquartile range

ICS: Immune confounder scale

CI: Confidence intervals

LC-MS/MS: Liquid chromatography with two mass spectrometry

HNMR: Proton nuclear magnetic resonance

HPLC: High-performance Liquid-Chromatography.

ELISA: Enzyme-Linked immunosorbent Assay
